# Genetic study of intrahepatic cholestasis of pregnancy in 101,023 Chinese women unveils East Asian-specific etiology linked to historic HBV infection

**DOI:** 10.1101/2024.07.01.24309754

**Authors:** Yanhong Liu, Yuandan Wei, Xiaohang Chen, Shujia Huang, Yuqin Gu, Zijing Yang, Liang Hu, Xinxin Guo, Hao Zheng, Mingxi Huang, Shangliang Chen, Tiantian Xiao, Yang Zhang, Guo-Bo Chen, Likuan Xiong, Xiu Qiu, Fengxiang Wei, Jianxin Zhen, Siyang Liu

## Abstract

**Background & Aims:** Intrahepatic cholestasis of pregnancy (ICP) is the most common and high-risk liver disorder during the critical period of human reproduction. Despite varying prevalence across populations, a mechanistic understanding of this phenomenon is lacking. This study delves into the genetic etiology of ICP in East Asians, drawing comparisons with Europeans to comprehend ICP etiology in the context of genetic background and evolution.

**Methods:** We conducted the hitherto largest-scale genome-wide association studies (GWAS) on total bile acid concentration (TBA) and ICP among 101,023 Chinese pregnancies. The findings were subsequently replicated in two cohorts and compared with European populations. Additionally, phenome-wide association and spatio-temporal evolution analyses were employed to understand the function and explore evolutionary pattern of sites associated with ICP.

**Results:** We identified eight TBA and five ICP loci, including ten novel loci. Notably, we found an East-Asian-specific genetic locus at 14q24.1, contributing to a 6.41 µmol/L increase in TBA and a 15.23-fold higher risk of ICP per risk allele (95% *CI*: 15.10 to 15.36, *P* = 9.23×10^-375^). Phenome-wide association studies and spatial-temporal evolution analyses revealed that the 14q24.1 ICP risk locus exhibits resistance to hepatitis B infection and has become prevalent only within the last 3,000 years in East and Southeast Asia.

**Conclusions:** Our investigations have unraveled a distinct etiology of ICP between Europeans and East Asians, and has linked ICP etiology in East Asians to a historical HBV epidemic in East and Southeast Asia within the last 3,000 years. These findings lay the groundwork for an improved biological understanding of ICP pathophysiology. Further exploration and utilization of these variations hold the potential for more precise detection, assessment, and treatment of ICP.

**Lay summary:** Intrahepatic cholestasis of pregnancy (ICP) is a prevalent and high-risk liver disorder that occurs during pregnancy, a critical period in human reproduction. It affects approximately 1% to 6.06% pregnancies and has been associated with severe adverse outcomes such as preterm birth and stillbirth. While rare and common variants associated with ICP have been identified in the European population, the genetic basis of ICP in East Asian population remains uncharacterized. Here, we conducted the largest-scale genome-wide association studies to date for TBA and ICP among 101,023 Chinese pregnant women, including 4,703 cases and 96,320 controls from two hospitals in Shenzhen, China. We replicated our findings in two independent Chinese cohorts and compared them with ICP genetic studies in the European population. We identified eight and five genome-wide significant loci for TBA and ICP, respectively, including ten novel loci. Notably, we identified an East-Asian-specific genetic locus contributing to a 6.41 µmol/L increase in TBA per risk allele and a 15.23-fold higher risk of ICP. Further exploration through phenome-wide association studies and spatial-temporal evolution analyses revealed that the 14q24.1 ICP risk locus exhibits resistance to hepatitis B infection and has become prevalent only within the last 3,000 years in East and Southeast Asia. These findings suggest a historical HBV epidemic in East and Southeast Asia within 3,000 years may have contributed to the increased prevalence of ICP and TBA risk alleles among East Asians. Our study unravels a distinct genetic etiology of ICP between Europeans and East Asians. These findings lay the foundation for an improved understanding of ICP pathophysiology and emphasize the need for integrating population evolution into genetic medicine for personalized genomics and clinical guidance.

**Highlights:**

(1) In the most powerful genome-wide association studies on TBA and ICP in East Asians to date, we identified eight and five genetic loci, respectively, of which, 7 and 3 were novel discoveries.
(2) One of the novel loci, the 14q24.1 locus, stands out as it contains unique causal genetic variants specific to East-Asians. These variants demonstrate large effects, contributing to an average increase of 6.41 µmol/L in TBA per risk allele and a 15.23-fold higher risk of ICP.
(3) The risk mutations associated with ICP at the 14q24.1 exhibit resistance to hepatitis B infection and has only become prevalent within the last 3000 years in East and Southeast Asia.

Abstract Figure:

Genetic basis and evolutionary history of intrahepatic cholestasis of pregnancy in East Asia.
TBA: Total bile Acid. ICP: Pregnancy intrahepatic cholestasis. Refer to the main text for the illustration.

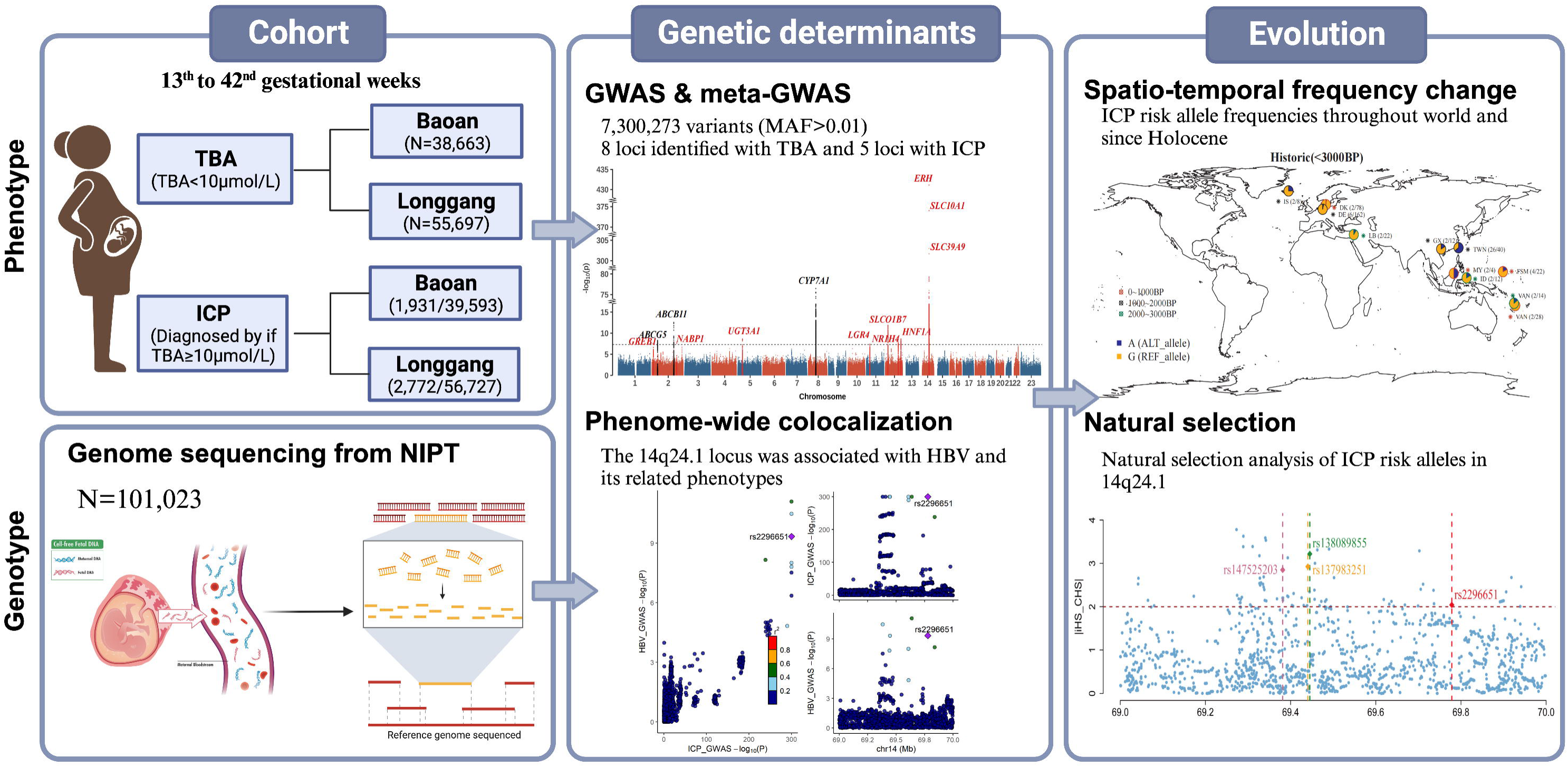

## Introduction

Intrahepatic cholestasis of pregnancy (ICP) is the most prevalent and high-risk liver disorder occurring during pregnancy, a critical phase in human reproduction^1^. The disease is characterized by maternal pruritus and elevated serum bile acid concentrations (≥10µmol/L), with diagnosis typically involving assessing maternal total serum bile acids (TBA) during routine pregnancy screening in the late second and third trimesters^2^. ICP has been associated with several severe adverse pregnancy outcomes, including increased risks of spontaneous and iatrogenic preterm birth, meconium-stained amniotic fluid, fetal asphyxia, and stillbirth^3, 4^. A large-scale meta-analysis of ICP cases revealed that a TBA level of 100 µmol/L is associated with an elevated risk of stillbirth, with a hazard ratio [*HR*] of 30.5 [95% *CI*: 8.83–105.30], compared to a TBA level below 40 µmol/L^5^. Several retrospective studies and case reports underscore the unpredictability of prenatal fetal death in patients with ICP after 36 weeks of pregnancy^2, 6, 7^. At present, oral ursodeoxycholic acid (UDCA) is the most commonly employed treatment in clinics to alleviate maternal pruritus and improve liver function. However, a recent clinical randomized controlled trial declared that treatment with UDCA does not reduce adverse perinatal outcomes in women with ICP^1^, leaving its effectiveness in enhancing perinatal outcomes and the long-term health of mothers with ICP and their children inconclusive^8^.

Unraveling new treatments of ICP necessitates a profound understanding of the disease’s etiology. Accumulated evidence suggests the complexity of ICP etiology, which is associated with genetic, endocrine, and environmental factors^9^. Familial clustering of ICP strongly implies a genetic basis^10^. Family-based studies indicate that the heterozygous state for an *MDR3* (alias *ABCB4*) gene defect likely represents a genetic predisposition within families^11, 12^. A candidate gene analysis study identified six SNPs in each of the *ABCB4* and *ABCB11* genes that demonstrated a significant association with ICP^13^. Case-control study also showed that *ABCB4*, *ABCB11,* and *ATP8B1* genes are related to intrahepatic cholestasis of pregnancy^14^. Case reports have proposed that NTCP deficiency, encoded by *SLC10A1* gene, might be a genetic factor contributing to ICP^15^. A recent genome-wide meta-association study involving 1,138 ICP cases and 153,642 controls from three Europeans studies identified eleven genes associated with ICP risk, most of which are related to hepatic functions^16^.

Notably, global prevalence of ICP varies substantially among different ethnicities. ICP affects approximately 0.32% of pregnancies in the United States, 5.6% in the Latina population in Los Angele^17, 18^, 0.7% in the United Kingdom^17^, 1.5%-4% in Chile^19, 20^ and 1.2% to 6.06% in China^21^. However, despite its variable geographical prevalence, factors influencing these differences are poorly understood. In addition, despite the considerably higher prevalence of the disease among East Asians, no studies of sufficient power have been conducted among the East Asian populations.

To address these gaps, we conducted the most powerful genome-wide association study (GWAS) of TBA and ICP in East Asia to date, utilizing sequencing data from the non-invasive prenatal testing (NIPT) of 101,023 Chinese pregnant women and comprehensive phenotypic records from two hospitals in Shenzhen city in South China. The discoveries were replicated in two independent Chinese datasets and compared to previous findings among the European population. Intriguingly, we found distinct etiology of ICP among Europeans and Asians. Among the ten novel loci previously not reported in the GWAS catalog, a genetic locus in 14q24.1 present exclusively among East Asians stands out, contributing to an average increase of 6.41 µmol/L in TBA per risk allele and a 15.23 times higher risk of ICP in East Asians. Further phenome-wide association and spatio-temporal evolutionary studies have linked this ICP risk locus to a putative HBV epidemic in East and Southeast Asia within the last 3000 years.

## Result

### Study design and TBA phenotypic distribution

The design of the study is summarized in **Abstract Figure**. In Shenzhen city, China, government-sponsored Non-Invasive Prenatal Tests (NIPT) were administered as a standard examination. Over the period from 2017 to 2022, we recruited 101,023 pregnant women during routine obstetric examinations across two Shenzhen hospitals (Baoan and Longgang). Each pregnant woman underwent both a NIPT test and at least one TBA test between the 13^th^ and 42^nd^ gestational weeks. After excluding outliners, 94,360 pregnancies with normal TBA levels (< 10µmol/L) were included for the GWAS of plasma TBA levels. Using the inclusion and exclusion criteria detailed in the Methods section, 4,703 pregnancies were identified as ICP cases, while 96,320 individuals served as controls for the ICP GWAS.

**Supplementary Fig. 1** illustrates a positively skewed phenotypic distribution of TBA levels, which exhibits a consistent median of 2.90 µmol/L (Interquartile Range [IQR]: [2.00, 4.20]) among normal pregnancies and a median of 14.52 µmol/L (IQR: [11.58, 23.10]) among ICP cases. The distribution of TBA levels, maternal age (29.72 ± 4.47 years old), and gestational week (33.05 ± 8.48 weeks) are similar between the two hospitals (**Supplementary Table 1)**.

We employed a well-established genetic analysis pipeline developed in our previous studies to analyze the NIPT data^22, 23^ and conduct the GWAS for TBA level and ICP across over 7 million sequence variants with minor allele frequency (MAF) greater than 1% in the study cohorts. We replicated the genetic findings in two independent Chinese datasets and provided meta-analysis estimates. We further conducted phenome-wide colocalization analyses to examine the genetic associations across a hundred pregnancy phenotypes. For the most significant and East Asian-specific 14q24.1 locus, we conducted spatio-temporal and natural selection evolutionary analyses to explore the frequency changes of the ICP risk alleles globally and since the Holocene.

### Genome-wide association study of TBA and ICP among the 101,023 pregnancies

Power estimations for the meta-GWAS design indicate an 80% statistical power to detect genetic associations with an effect size (*β*) > 0.170 and MAF ≥ 0.05, or otherwise *β* > 0.370 and MAF ≥ 0.01 for TBA, and similarly for genetic associations with an odds ratio (*OR*) > 1.140 and MAF ≥ 0.05 as well as an *OR* > 1.320 and MAF ≥ 0.01 for ICP **(Fig. S2)**. The genomic inflation (lambda) values for TBA and ICP were 1.069 and 1.018, respectively, suggesting negligible confounding of population stratification **(Fig. S3)**.

In total, we identified 1,673 and 1,281 genome-wide significant variants (*P* < 5×10^-8^), constituting 19 and 16 independent association signals for TBA and ICP respectively (**Table S2**). These were represented by 8 independent association loci for TBA and 5 for ICP **(Fig. 1**; **Table 1; Table S3)**. Among the 13 association loci, 7 and 3 were not previously reported in the GWAS catalog or Phenoscanner and were identified as novel loci (gene symbols in red in **Fig. 1**).

**Fig. 1.**
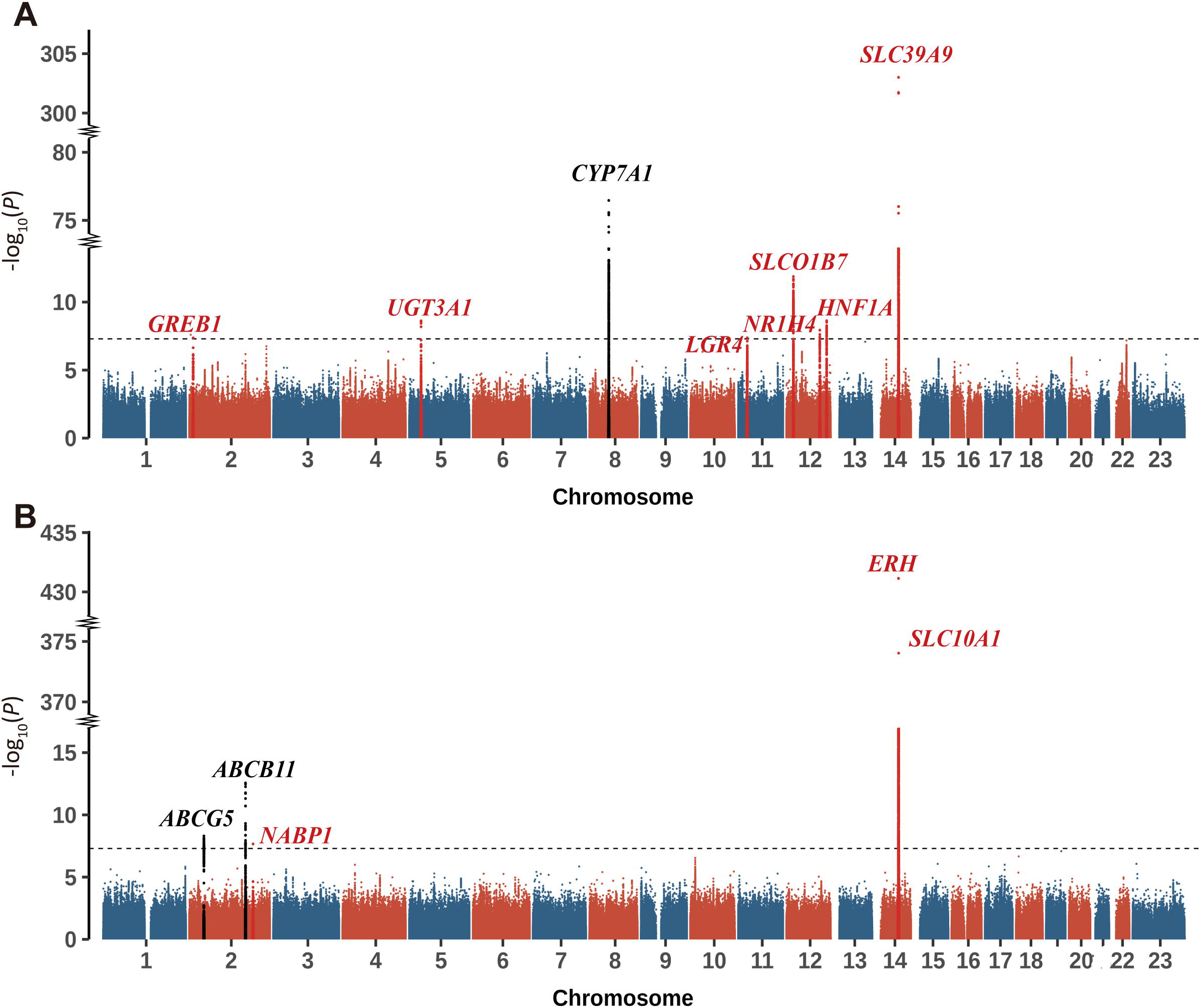
Meta-analysis of Genome-wide association study for total bile acid (TBA) and intrahepatic cholestasis of pregnancy (ICP). (A) Manhattan plot for TBA GWAS meta-analysis, encompassing 94,360 samples. (B) Manhattan plot for ICP GWAS meta-analysis, involving 4,703 cases and 96,320 controls. Chromosomes are ordered on the x-axis and the -log_10_(*P*) values for the association tests are shown on the y-axis. Horizontal dashed lines delineate the genome-wide significance threshold (*P* = 5×10^-8^, in grey). Eight and five independent loci achieved genome-wide significance (*P* < 5×10^-8^) with TBA and ICP, respectively. Labels in black denote known loci and labels in red highlight novel loci.

We assessed the fidelity of the association signals by comparing the effect estimates between the two independent hospitals (internal replication) and with two external Chinese datasets (external replication) (**Materials and Methods**). The effect estimates demonstrated high consistency between the two hospitals, where 18/19 association signals (94.8%) or 7/8 loci (87.5%) exhibited the same beta direction and surpassed Bonferroni correction significance levels for TBA in each hospital. Similarly, for ICP, all 16 association signals included in the 5 loci exhibited the same effect direction and significant *P* value after Bonferroni-correction (**Fig. S4, Table S2-S3**). The only *GREB1* gene locus (lead SNP rs10929754-T) associated with TBA did not pass the Bonferroni correction significance level in the Baoan cohort. However, it consistently showed the same effect direction and achieved nominal significance (*P*=0.0233). The effect estimates also demonstrated high consistency between our study and the two Chinese external datasets, where all signals and loci were replicated in one of the two external datasets **(Fig. S5, Table S2, Table S4)**. The LocusZoom regional association maps for the eight loci associated with TBA and the five loci associated with ICP are presented in **Fig. S6-S7.** The above evaluations suggest high fidelity of the associations identified in our study.

### The East Asian-specific at 14q24.1 locus contributes to a 15.23-fold higher risk of ICP

Seven out of the eight loci associated with TBA and three of the five loci associated with ICP were not previously documented in the GWAS catalog^24^ and Phenoscanner^25^, particularly in the current unique ICP GWAS among the European population^16^. Genes contained within the genetic loci associated with TBA levels and ICP are enriched in bile acid and bile salt transport, bile acid signaling and biosynthetic processes, as well as lipid metabolism (**Table S5**). Specifically, the *ABCB11*, *ABCG5* and *SLC10A1* loci associated with ICP are uniquely expressed in liver tissue^26^ and hepatocyte cells^27^, reflecting the central role of the liver organ in the development of ICP. Notably, we observed substantial discrepancies in genetic discoveries between our study and the European ICP GWAS study (**Fig. S8, Table S2 and Table S5**). Particularly, variants within the 14q24.1 locus, encompassing the *ERH*, *SLC39A9* and *SLC10A1* genes, are absent outside East Asian populations. This includes the lead SNPs rs137983251-G associated with TBA, the lead SNP rs147525203-C associated with ICP and the missense variants rs2296651-A (*p.Ser267Phe*) present in *SLC10A1* which is strongly associated with TBA and ICP risk (*MAF*=0.04, *OR*=15.23, [95%*CI*]: 15.23 [15.10, 15.36], *P* = 9.23×10^-375^). While rs137983251-G and rs147525203-C are in complete linkage disequilibrium (LD) with each other (*R*^2^=1), rs2296651-A are in less strong LD with rs137983251-G and rs147525203-C (*R*^2^=0.21)^28^.

### The ICP risk variants at the 14q24.1 locus decrease HBV infection

Notably, *SLC10A1*, also known as *NTCP*, is recognized as a sodium/bile acid cotransporter and has been implicated as the cellular receptor for HBV infection^29^. Despite this, none of the GWAS studies cataloged in the GWAS catalog, Phenoscanner or PubMed has linked it to TBA or ICP^24, 25^, likely due to its population specificity. To elucidate the function of the East Asian-specific 14q24.1 ICP risk alleles, we scrutinized 246 phenotypes in the Biobank of Japan (BBJ)^30^ and over 100 pregnancy phenotypes in our dataset (see Materials and Methods for URL). We found that lead SNPs of TBA and ICP (rs137983251 and rs147525023) and the rs2296651-A (*p.Ser267Phe*) variant were significantly associated with the traits related to Hepatitis B virus (HBV) infection. In our GWAS of six HBV traits, the 14q24.1 locus stands out as the second most significantly associated locus with HBV infection, exhibiting strong associations with Hepatitis B surface antigen (HBsAg), Hepatitis B e antibody (HBeAb) and Hepatitis B core antibodies (HBcAb) (*P*<5×10^-8^) (**Fig. S9**). Stacked LocusZoom plots further confirmed the shared genetic effect between TBA/ICP, and HBV/HBsAg/HBeAb/HBcAb in the 14q24.1 locus **(Fig. S10)**. Other traits affected by the 14q24.1 locus include Alanine transaminase level during pregnancy (**Materials and Methods** for URL).

To fine map the potential causal variants contributing to both TBA/ICP and HBV infection, we conducted a colocalization analysis of TBA & ICP and six HBV-related traits on a 1Mbp region (chr14:69-70Mbp) consisting of 1,909 genetic variants (MAF>0.01). We found that TBA and ICP were both colocalized with HBV with at least one variant overlapping in their 95% credible set (PP.H4>0.75 & H4/H3>3) (**Fig. 2A-B, Fig. S11, Table S7)**. Evidence for the strongest colocalization of GWAS signals between TBA and HBV was found for *SLC39A9* (rs138089855, intron variant), and that between ICP and HBV was found for *SCL10A1* (rs2296651, *p.Ser267Phe*, missense variant). The rs138089855-C mutation and rs2296651-A mutation significantly affect immune evasion of HBV infection (*OR* ranging from 0.57 to 0.82, *P* < 4.63×10^-10^), reducing risks of HBsAg (*OR* ranges from 0.60 to 0.83, *P* < 3.72×10^-5^), HBeAb (OR ranges from 0.65 to 0.82, *P* < 3.24×10^-7^) and HBcAb (*OR* ranges from 0.63 to 0.79, *P* < 2.24×10^-11^) (**Fig. 2C-D**).

**Fig. 2.**
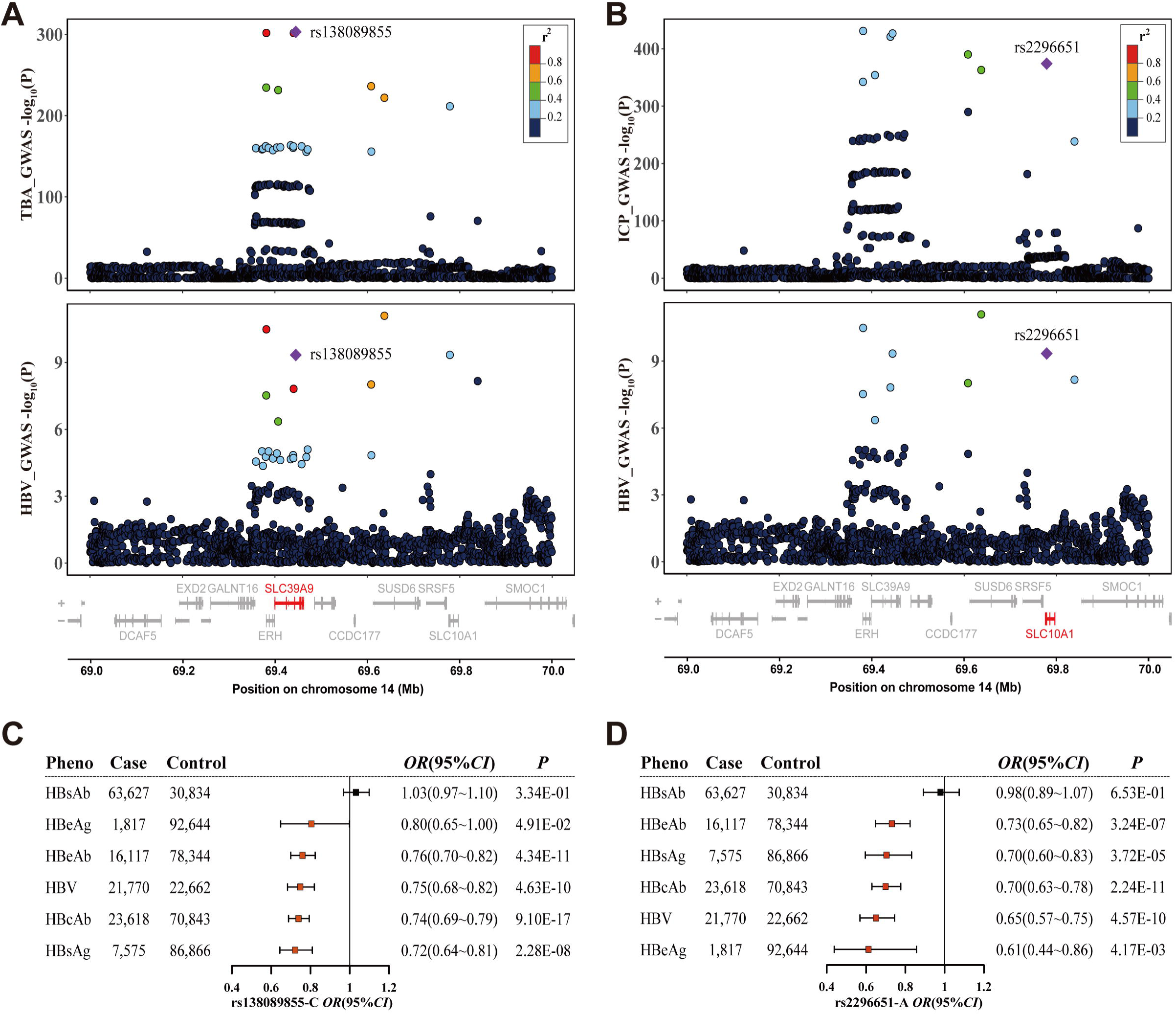
GWAS-GWAS colocalization of TBA & ICP with HBV and forest plots for the shared SNPs with HBV-related traits. (A) The purple diamond in the plot indicates the colocalized SNP, rs138089855, shared between TBA and HBV. (B) Another SNP, rs2296651, is identified as the colocalized SNP between ICP and HBV. The color coding on the plot signifies the *R*^2^ measure of linkage disequilibrium of the colocalized SNP. The forest plot illustrates the genetic effect of (C) rs138089855-C allele and (D) rs2296651-A allele at 14q24.1 on the meta-analysis for HBV and five HBV antigen & and antibody status. The error bars represent the 95% confidence interval of the odds ratio (*OR*).

### The *p.Ser267Phe* mutation of the *SLC10A1* variant became prevalent within the last 3,000 years in the areas of East and Southeast Asia

The Geography of Genetic Variants Browser (https://popgen.uchicago.edu/ggv/) illustrates that the lead SNP of TBA (rs137983251-G) and ICP (rs147525203-G), as well as the rs138089855-C and missense locus rs2296651-A identified in colocalization analysis, are only present in East and Southeast Asia **(Fig. S12)**. To delve further into the historical evolutionary history of ICP genetic loci, we assessed the spatio-temporal frequency differences of rs2296651-A globally and since the Holocene using the Allen Ancient Data Resource (AADR)^31^. In the Holocene era (10,000 BP before now), only one sample with the *p.Ser267Phe* mutation exists in Morocco (**Fig. 3A, Table S8**). In the Neolithic era (10,000∼3,000BP), several *p.Ser267Phe* variations appear sporadically, with single samples distributed in Italy, Russia, Spain and California, USA (**Fig. 3B, Table S9**). Notably, until the Historic (<3,000BP), a large number of mutations occurred at this locus, especially within the range of 1,000 to 2,000BP (**Fig. 3C, Table S10**). Currently, the ICP risk mutations of this locus are only distributed in East Asia and Southeast Asia (**Fig. 3D, Table S11)**.

**Fig. 3.**
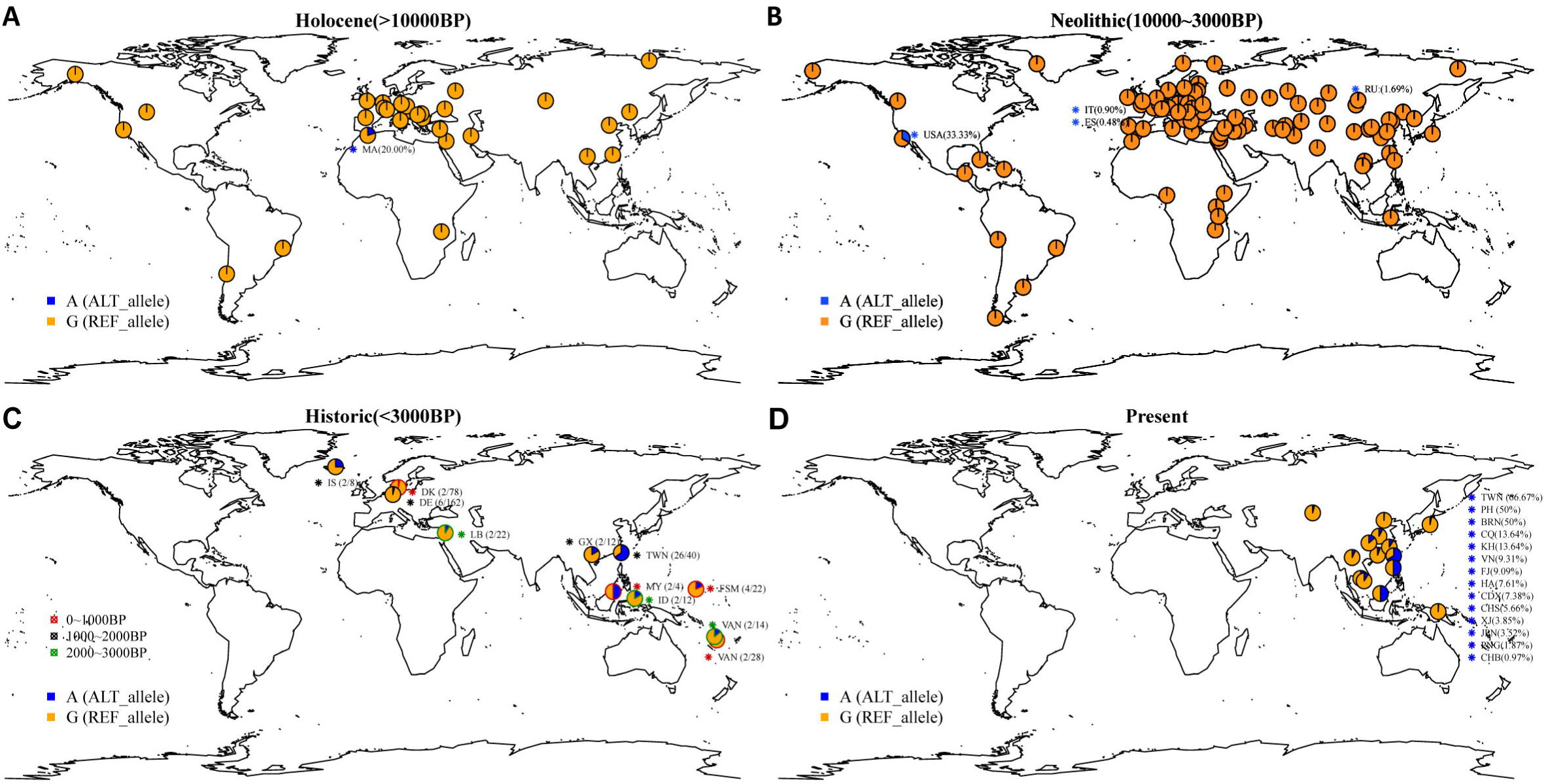
Temporal and geographical changes of allele frequency of rs2296651. The geographical frequency distribution of SNP rs2296651 is depicted in (A) the Holocene age (> 10,000BP), (B) the Neolithic age (10,000∼3,000BP), (C) the Historic (<3,000BP), and (D) the present-day global populations, utilizing the “1240K” dataset (“Allen Ancient DNA Resource”, version 54.1). The ancestral allele (reference allele) rs2296651-G is represented in orange and the derived allele (alternative allele A, the ICP risk allele) is shown in blue. The color of the pie chart border indicates the detailed archaeological period in the plot in (C) (i.e., red: 0∼1,000BP, black: 1,000∼2,000BP, green: 2,000∼3,000BP). Plots (C) and (D) exclusively display populations where rs2296651-A is present. The frequency or quantity of the A allele in different populations is shown in parentheses. Detailed allele frequencies are provided in **Supplementary Table 8-11**.

### Natural selection signals in the 14q24.1 locus

The distinctive prevalence of the 14q24.1 locus in different geographical regions may indicate natural selection. To further understand the genetic diversity in the 14q24.1 locus, we employed 1KGP high-depth sequencing data for selection analysis. We calculated several metrics based on the site-frequency spectrum in this region for three representative populations: Southern Han Chinese (CHS), Northern Europeans from Utah (CEU), and Yoruba in Ibadan, Nigeria (YRI). We observed a decrease in nucleotide diversity in chr14:69.0-69.2Mbp and around chr14:69.8-69.9 regions in CHS compared to CEU and YRI. Tajima’s *D* and Fay and Wu’s *F* statistics also suggested an excess of high-frequency derived SNPs in these two regions **(Fig. S13)**.

Furthermore, we conducted a selection analysis based on population differentiation and linkage disequilibrium. The chr14:69.0-69.2Mbp region shows the highest locus-specific branch length (LBSL) statistics for CHS compared with the CEU using AFR as the reference panel, indicating a signal of positive selection in East Asia. The results of iHS and the negative XP-EHH values also support that the selection occurred in the CHS population **(Fig. S14)**.

Regarding the high linkage between lead SNPs of TBA and ICP, we explored whether there are differences in haplotype frequencies among different races. We extracted SNPs at high linkage disequilibrium with SNP rs147525203, rs137983251, rs138089855, or rs2296651, resulting in 16 SNPs which were phased into haplotypes. Having examined differences between different races, we found significant differences in haplotypes in East Asians compared to other populations. Of the top 10 haplotypes, 4 types (H4, H5, H6, and H9) were almost exclusively found in 10.8% of Asian individuals **(Fig. S15, Table S12)**. The origin of these haplotypes was not linked to known archaic ancestry. However, we cannot be ruled out that these haplotypes may have originated from unknown East and Southeast Asian archaic hominins, which are currently underrepresented in human genetic investigations^32,33,34^.

## Discussion

Pregnancy constitutes a critical period for human reproduction, and ICP emerges as a high-risk liver disorder during this phase. Characterized by maternal pruritus and elevated serum bile acid concentrations^2^, ICP is associated with an elevated risk of severed pregnancy outcomes, including intrauterine fetal growth restriction, preterm birth, fetal asphyxia, still birth and lower birth weight^3, 4^. Standard clinical practice typically designates pregnancies diagnosed with ICP as high-risk, administering UDCA post-diagnosis, and often inducing labor toward full term. However, recent evidence from an RCT challenges the efficacy of UDCA in reducing adverse perinatal outcomes in women with ICP^1^. The high risk and lack of effective treatment of ICP underscores the need for a more comprehensive understanding of the factors leading to ICP. In the present study, we conducted the first-ever GWAS meta-analysis of TBA and ICP traits in East Asians, encompassing 101,023 participants, including 4,703 cases and 96,320 controls, with signals verified in two independent Chinese cohorts. We identified eight and three genome-wide significant associated loci for TBA and ICP, respectively, including seven and three novel association loci.

Of particular interest is the revelation that the most significant 14q24.1 locus contributes to an average odds ratio of 15.23 for ICP in Chinese pregnancies. This is much higher compared to the 1.70 odds ratio for ICP associated with the known *ABCG5* and *ABCB11* loci reported in the European population. The high-risk allele in the 14q24.1 locus, comprising the *SLC39A9* (lead SNP rs137983251), *ERH* (lead SNP rs147525023), and *SLC10A1* (missense variant rs2296651-A, *p.Ser267Phe*) genes, is uniquely present in the East and Southeast Asia and conspicuously absent in other global populations. Comprehensive phenome-wide association scans based on BBJ data and our own dataset revealed a significant association between the ICP-high-risk allele and a protective effect against HBV infection, reflected by a decreased risk of HBV infection, HBsAg, HBcAb, and HBeAb serological positivity. Spatiotemporal and natural selection analyses further suggest that the prevalence of the ICP risk allele dates back within the last 3000 years, putatively due to selection over resistance of HBV infection.

Currently, Hepatitis B virus (HBV) infection remains a significant public health concern, affecting over 296 million people worldwide chronically infected^35^. However, little is known about the origin and historical spread of HBV. In 1964, American physician and geneticist Blumberg et al discovered a new antigen in the serum of an Australian Aborigine named Australian antigen (AuAg)^36^, and subsequently identified it as the surface antigen (HBsAg), marking the first specific indicator of hepatitis B virus (HBV)^37^. Recent evidence of HBV’s historical existence extends to mummies in Korea and Italy from 400 years ago^38, 39^. Moreover, in 2018, the discovery of HBV genomes from ancient DNA derived from human skeletal remains fossilized in the Neolithic period around 7,000 years ago was documented^40^. Our study suggests that a potential HBV epidemic occurred 3,000 years ago in East and Southeast Asia, leading to a balancing or positive selection of the ICP risk allele. This selection resulted in a higher prevalence of ICP observed currently in these regions, shaping a distinct genetic etiology of ICP between East Asians and Europeans.

What are the medical implications arising from these findings? The first takeaway is that the genetic etiology of ICP differ substantially between Asian and European populations. Clinical practice should take into account the ethnic ancestry of patients for informed clinical decisions, with particular attention to the presence of the 14q24.1 ICP risk allele. Secondly, the prevalence of ICP in East and Southeast Asia is an outcome of evolutionary processes. Pregnant individuals carrying the risk alleles exhibited enhanced biological resistance to HBV infection and are likely descendants of survivors from a historical HBV epidemic. Mothers carrying the risk allele need not bear any stigma regarding their infants. Thirdly, evidence on the clinical risk of ICP primarily stems from large-scale cohort-based observational studies^5^, susceptible to confounding by unknown factors. Given the cumulative substantial impact of TBA and ICP risk alleles identified in this study, more refined causal inference regarding the relationship between TBA and ICP and the short-term birth outcomes and long-term offspring health, can be achieved using methods such as Mendelian randomization applied to East-Asian birth cohorts^41, 42^.

Building upon these discoveries, more precise clinical decisions can be tailored for pregnancies, preventing unnecessary over-treatment. Additionally, novel therapeutics can be developed specifically for high-risk patients based on their genetic origins, compensating for the limited effectiveness of UDCA in reducing adverse outcomes for ICP patients. For example, as HBV vaccinations are mandatorily implemented for newborns in countries like China, it may be worthwhile to explore whether increasing NTCP activity may benefit the ICP patients in clinics while not increasing their risk for HBV infection.

## Materials and Methods

### Cohort description

The study encompassed 70,608 pregnancies recruited from Longgang District Maternity & Child Healthcare Hospital of Shenzhen City (Longgang cohort) and 50,948 pregnancies from Shenzhen Baoan Women’s and Children’s Hospital (Baoan cohort). These participants engaged in a routine pregnancy screening program in Shenzhen between 2017 and 2022. Pregnancies involving multiple gestations and those lacking TBA level measurements were excluded. Finally, our study comprised 101,023 pregnant women who underwent at least one TBA level assessment during gestational weeks 13 to 42.

To conduct GWAS of TBA and ICP, we integrated NIPT data with clinical phenotypic data (see **Supplementary Methods**). All participants provided written informed consent. The study received approved from the Medical Ethics Committee of the School of Public Health (Shenzhen), Sun Yat-sen University (No. 2022-021), Longgang District Maternity and Child Healthcare Hospital of Shenzhen City (LGFYYXLLL-2022-024), and Shenzhen Baoan Women’s and Children’s Hospital (LLSC-2021-04-01-10-KS). Data collection was also approved by the Human Genetic Resources Administration of China (HGRAC) (Baoan cohort: [2023]CJ1415, Longgang cohort:[2023]CJ1455).

### Phenotype definition

The TBA level was defined as the peak level observed for each individual during gestational weeks 13 to 42 after removing outliers (N = 94,360). ICP cases were identified as pregnancies with TBA concentrations ≥ 10µmol/L during the same gestational period. In total, 4,703 cases and 96,320 controls for ICP were included in the study.

We also obtained the hepatitis B antigen and antibody measurements from data of the pregnant screening program. The phenotype of hepatitis B virus (HBV) infection was defined based on hepatitis B antigen and antibody. Detailed information can be found in **Supplementary Methods**.

### Genome-wide association analysis

We conducted the GWAS analysis using PLINK 2.0^43^. Covariates such as gestational week, maternal age, and the top ten principal components accounting for population stratification were included in the analysis. The quantitative phenotype (TBA) underwent a rank-based transformation to achieve a normal distribution.

To integrate results from two cohorts, a meta-analysis was performed using fixed-effect models with inverse-variance weighting, employing METAL software (version 2011-03-25)^44^. Variants with a minor allele frequency (MAF) less than 0.01 were excluded. Conditional and joint analysis (GCTA-COJO) was performed using GCTA software^45, 46^ to identify independent genome-wide significant signals. A stepwise model selection procedure (--cojo-slct) with a collinearity threshold of 0.2 was employed to choose independently associated SNPs, with the reference panel from the Born in Guangzhou Birth cohort (BIGCS)^42^ for the LD structure of the variants. Subsequently, we delineated the range of 500kb upstream and downstream blocks of a significant signal into separate locus, considering the SNP with the lowest *P* value as the lead SNP.

### Statistic power calculation

All statistical analyses were executed in R (version 4.2.1). Post hoc power calculations were performed to access the spectrum of effect size (*β*)/odds ratio (*OR*) and allele frequencies at which associations could be detected at genome-wide significance (*P* < 5×10^-8^) within a specified number of samples/cases and controls in the meta-analysis. The calculation was performed using the R package “genpwr”^47^ (version 1.0.4) using a linear or logistic model under a genetic additive mode.

### Replication and comparison

For internal replication, we applied stringent criteria to define a locus as replicated, requiring the same beta direction and *P* values reaching the Bonferroni correction threshold in both the Baoan and Longgang cohorts. In the external replication, we replicated genome-wide significant SNPs using two independent study cohorts (Baoan NIPT PLUS cohort and BIGCS cohort). SNPs meeting the following criteria were regarded as replicated: 1) they exhibited a consistent direction of effect for lead SNPs between the discovery and the replication cohorts and 2) they reached Bonferroni-corrected *P* values or passed a two-sided two-sample t-test. Further details on external replication can be found in **Supplementary methods.**

Furthermore, we compared the genetic influence of SNPs with a previously published European cohort^16^. This comparison involved assessing the direction of genetic effects and Bonferroni-corrected *P* values of lead SNPs between our meta-analysis results and the European cohort.

### PheWAS and Colocalization analysis

A PheWAS was conducted using 246 phenotypes from BBJ (https://pheweb.jp/) and 131 pregnancy phenotypes from the MONN PheWeb established by our research group (http://47.112.105.165/). Regions showing evidence of colocalization between the GWAS and GWAS signals were identified utilizing pre-defined thresholds: PP4 (posterior probability that there exists a single causal variant common to both traits) ≥ 0.75 and PP4/PP3 ≥ 3, employing the R package “coloc” (version 5.1.0.1)^48^. The prior probabilities for SNP association with either of the two traits or with both traits were set to 1 × 10^−4^ and 1 × 10^−5^, the default values, respectively.

### Geographical frequency distribution analysis

To examine the frequency distribution of identified SNPs across various ancient age groups, we employed ancient DNA data from the compiled 1240K dataset sourced from the Allen Ancient DNA Resource (version 54.1)^31^. The geographical map, supplemented with pie charts, was created using R packages “maps” and “mapplots” to illustrate the geographical frequency distribution.

### Modern DNA-based selection test

We performed three types of selection tests for modern DNA, encompassing analyses based on site-frequency spectrum (SFS), population differentiation, and haplotype selection to explore signals of positive or balancing selection within East Asia populations. SNPs with minor allele frequencies (MAF) < 0.01 in all five super-populations (AFR, CEU, SAS, EAS and AMR) and genetic variants lacking ancestral information were excluded. Ultimately, 18,221,282 SNPs were used for the natural selection analysis (**Supplementary Methods** for details).

### Haplotype analysis

The data used for haplotype analysis corresponds to that employed in the aforementioned natural selection analysis. SNPs with an MAF < 0.05 in all 5 super-populations and genetic variants lacking ancestral information were excluded. We extracted SNPs exhibiting high linkage disequilibrium (*R^2^*> 0.20) with rs137983251, rs138089855, rs147525203, or rs2296651 variants, resulting in 16 SNPs that were used to construct 31 haplotype types. To further streamline haplotype diversity, the rs79422091 variation was excluded to merge similar haplotypes. Consequently, 16 SNPs were selected to form 25 distinct haplotypes.

## Abbreviations

AADR: Allen Ancient DNA Resource
BRN: Brunei
CDX: Chinese Dai in Xishuangbanna
CHB: Han Chinese in Beijing
CHS: Southern Han Chinese
CI: confidence interval
COJO: conditional and joint analysis
CQ: Chongqing (China)
DE: Germany
DK: Denmark
ES: Spain
FJ: Fujian (China)
FSM: Federated States of Micronesia
GWAS: genome-wide association study
GX: Guangxi (China)
HA: Henan (China)
HBcAb: hepatitis B core antibody
HBeAb: hepatitis B e antibody
HBeAg: hepatitis B e antigen
HBsAb: hepatitis B surface antibody
HBsAg: hepatitis B surface antigen
HBV: hepatitis B virus
HCV: hepatitis C virus
HR: hazard ratio
ICP: intrahepatic cholestasis of pregnancy
ID: Indonesia
iHS: integrated Haplotype Score
IS: Iceland
IT: Italy
JPN: Japan
KH: Cambodia
LB: Lebanon
LD: linkage disequilibrium
MA: Morocco
MAF: minor allele frequency
MY: Malaysia
NIPT: non-invasive prenatal testing
OR: odds ratio
PCs: principal components
PH: Philippines
PNG: Papua New Guinea
PP.H: posterior probability for hypothesis
RU: Russia
SFS: site-frequency spectrum
SNP: single nucleotide polymorphism
TBA: total serum bile acids
TWN: Taiwan (China)
USA: United States of America
VAN: Vanuatu
VEP: Ensembl Variant Effect Predictor
VN: Vietnam
XJ: Xinjiang (China)
XP-EHH: Cross Population Extended Haplotype Homozygosity.

## Data availability

GWAS summary statistics for TBA and ICP phenotypes will be made publicly available in the GWAS catalog (https://www.ebi.ac.uk/gwas/) upon publication.

## Author contribution

SL, FW, JZ, and XQ conceived the study. JZ, XC, LH, YW, ZY, HZ, SC, and TX collected and organized the data from the maternity testing system. YL, YG, XC, LH, and XG conducted data pre-processing and preliminary analyses. YL performed all statistics, GWAS, and evolutionary analysis. YL and YW performed the visualization of all results. JZ, SH, MH, and XQ provided the validation data. SL, FW, JZ, XQ, YZ, GBC, and LX provided professional guidance and interpretation of data. YL & SL wrote the manuscript with input from all authors. All authors contributed to manuscript revisions and approved the final version of the article. SL is responsible for the integrity of the work as a whole.

