## Supplementary material for "Genetic study of intrahepatic cholestasis of pregnancy in 101,023 Chinese women unveils East Asian-specific etiology linked to historic HBV infection": Table 1

**Tables**

**Table 1. Genome-wide significant loci of TBA & ICP.**

| **CHR:BP** | **SNP** | **A1** | **A2** | **GENE** | ***EAF*** | ***BETA*** | ***SE*** | ***P-value*** |
| --- | --- | --- | --- | --- | --- | --- | --- | --- |
| ***TBA(μmol/L)*** | | | | | | | | |
| **2:11542948** | rs10929754 | T | G | *GREB1* | 0.2630 | -0.04 | 0.01 | **4.26E-08** |
| **5:35975464** | rs6861226 | G | A | *UGT3A1* | 0.4697 | -0.03 | 0.01 | **2.77E-09** |
| **8:58479765** | rs9297994 | G | A | *CYP7A1* | 0.2044 | -0.11 | 0.01 | **3.45E-77** |
| **11:27369903** | rs3782109 | C | T | *LGR4* | 0.4814 | 0.03 | 0.01 | **4.22E-08** |
| **12:20981503** | rs11835351 | A | G | *SLCO1B7* | 0.0629 | 0.07 | 0.01 | **1.33E-12** |
| **12:100506665** | rs11110399 | A | G | *NR1H4* | 0.2922 | -0.03 | 0.01 | **1.15E-08** |
| **12:120986687** | rs2243458 | T | C | *HNF1A* | 0.4024 | 0.03 | 0.01 | **2.35E-09** |
| **14:69440428** | rs137983251 | G | A | *SLC39A9* | 0.0238 | 0.55 | 0.01 | **2.13E-302** |
| ***ICP*** | | | | | | | | |
| **2:43833693** | rs6716451 | T | C | *ABCG5* | 0.1637 | 0.20 | 0.03 | **4.97E-09** |
| **2:169003206** | rs112779928 | T | C | *ABCB11* | 0.3499 | -0.23 | 0.03 | **2.74E-13** |
| **2:191584285** | rs4488606 | A | G | *NABP1* | 0.4442 | 0.25 | 0.05 | **2.17E-08** |
| **14:69381407** | rs147525203 | C | T | *ERH* | 0.0238 | 1.93 | 0.04 | **7.29E-432** |
| **14:69778476** | rs2296651 | A | G | *SLC10A1* | 0.0376 | 2.72 | 0.07 | **9.23E-375** |

CHR, chromosome; BP, base pair position; SNP, single-nucleotide polymorphism; A1, effect allele; A2, non-effect allele; EAF, effect allele frequency; OR, odds ratio; SE, stander error.
