## supplementary_information for "Genetic study of intrahepatic cholestasis of pregnancy in 101,023 Chinese women unveils East Asian-specific etiology linked to historic HBV infection"

**Supplementary Fig. 13. Site Frequency Spectrum-based selection tests with modern DNA samples from 1000 Genomes Project across three populations. 36**

### Supplementary Methods

#### GWAS study design and cohort description

Previous study has pointed that the more accuracy for presenting population genetic variation and inferencing population structure is achieved with very large sample sizes, even when utilizing extremely low sequencing depth[1]. Furthermore, previous study has demonstrated the substantial value of non-invasive prenatal testing (NIPT) data for GWAS analysis[2].

A total of 121,556 Chinese pregnancies who underwent non-invasive prenatal testing (NIPT) at two hospital cohorts in Shenzhen, China, were enrolled in our study between 2017 and 2022. The Baoan study cohort included 70,608 pregnant women recruited at the Shenzhen Baoan Women's and Children's Healthcare Hospital (Shenzhen, China). Additionally, 50,948 pregnant women who visited Shenzhen Longgang District Maternity & Child Healthcare Hospital (Shenzhen, China) were recruited for the Longgang study cohort. Here, we excluded 5,625 pregnancies potentially involving multiple gestations, along with 14,908 pregnancies for which TBA concentration results were unavailable during this period.

Using the method described in the previous study[2], maternal genotypes were inferred from NIPT ultra-low-depth whole-genome sequencing data. Phenotypic data were obtained from the hospital's electronic medical system during routine pregnancy screening program. Subsequently, we combined genotype data with clinical phenotypic data to explore the genetic molecular basis of TBA and ICP.

This study was reviewed and approved by Ethics Committee of School of Public Health (Shenzhen), Sun Yat-Sen University (2021. No.8), as well as the Institutional Board of Shenzhen Baoan Women's and Children's Hospital (LLSC2021-04-01-10-KS) and Longgang District Maternity and Child Healthcare Hospital of Shenzhen City (LGFYYXLLL-2022-024). The study strictly adhered to regulations governing ethical considerations and personal data protection. Data collection was approved by the Human Genetic Resources Administration of China (HGRAC). Written informed consent of all participants were obtained.

**Phenotype definition**

During the GWAS analysis, Hepatic biochemistry test data, including total bile acid concentrations, were obtained during second and third trimester pregnancy (13-42 gestational weeks) as part of routine pregnant screening in two hospital cohorts. The peak level for each individual during this period was defined as TBA quantitative trait (N = 94,360). ICP cases were identified as pregnancy with TBA concentrations ≥ 10μmol/L during the same period. In total, 4,703 cases and 96,320 controls for ICP were included in the study. The characteristics of the participants are presented in **Table S1**.

We also obtained the infection markers, including hepatitis B surface antigen (HBsAg), hepatitis B surface antibody (HBsAb), hepatitis B e antigen (HBeAg), hepatitis B e antibody (HBeAb) and hepatitis B core antibody (HBcAb), from pregnant screening. The sample size for HBsAg is 44,432 (7,575 cases versus 86866 controls), while the sample sizes for the other four traits are all 94,441 each (cases verus controls are 63627/30834, 1817/92644, 16117/78344, and 23618/70843, respectively). HBV persistent carriers and spontaneously recovered subjects were categorized as the HBV cases. HBV persistent carriers were defined as individuals positive for both HBsAg and HBcAb but negative for hepatitis C virus (HCV) antibody (anti-HCV). Spontaneously recovered subjects were those who were negative for HBsAg and anti-HCV but positive for both HBsAb and HBcAb. HBV controls were individuals negative for all HBV markers, including HBsAg, HBsAb, HBeAg, HBeAb, and HBcAb, as well as negative for HCV. In total, 21,770 cases and 22,662 controls of HBV were recruited in the study.

**Sequencing, alignment and imputation**

We collected sequencing data from non-invasive prenatal testing in both the Baoan and Longgang cohorts. The sequencing protocol details were outlined in Zhang et al[3] & Liu et al[2]. In summary, each participant underwent whole-genome sequencing, resulting in 9.9-21.9 million cleaned reads, corresponding to sequencing depths of approximately 0.11x-0.25x, with an average of 0.17x. Next, the single-end read alignment option in BWA was used to align the cleaned reads to the hg38 human genome reference[4]. The rmdup option in samtools was used to remove potential PCR duplicates[5]. The realign and base quality recalibration method in GATK were used to realign the reads and to recalibrate base quality score[6]. Finally, the alignment files were stored as bam files. After alignment, we employed GLIMPSE[7] (version 1.1.1) to impute genotype probabilities for all 121,556 individuals with a 10k Chinese reference panel. All alignment and imputation process were conducted at the National Supercomputing Center in GuangZhou.

**Genome-wide association analysis**

The GWAS analysis employed the multiple linear regression model for TBA and the logistic regression model for ICP to examine the association of SNPs using PLINK2.0 (https://www.cog-genomics.org/plink/2.0/). Gestational week, maternal age and the top ten principal components accounting for population stratification were included as covariates for TBA and ICP. Otherwise, we also conducted GWAS using logistic regression model for HBV and its antigen & antibody, with maternal age and the top ten principal components included as covariates. The additive genetic model of SNP dosage was utilized for genetic-phenotypic association. Principal component analysis (PCA) was conducted using PLINK2.0[8] on the dataset of 121,556 individuals.

The results of meta-GWAS were visualized with Manhattan plot using the R package ggplot2 (version 3.4.2) (https://cran.r-project.org/web/packages/ggplot2/index.html) and ggbreak (version 0.1.2) (https://cran.r-project.org/web/packages/ggbreak/index.html)[9]. Quantile-quantile (QQ) plots were generated using the observed and expected -log10 (*P* value) with R package ggplot2. The genomic inflation factor (lambda), calculated based on the 50th percentile, was 1.069 for TBA and 1.018 for ICP separately, indicating no significant population stratification. Regional high-resolution association plots showing the LD between markers in the lead loci were generated using LocusZoom (version 1.4) (http://locuszoom.org/).

**Variants Annotation**

Gene annotation was conducted using the Ensembl Variant Effect Predictor[10] (VEP, version 101), with indexed GRCh38 cache files (version 109). All the data utilized for annotation were obtained from the Ensembl FTP server (https://ftp.ensembl.org/pub/). Based on a set of VEP default criteria, the "--pick" option was used to assign a single consequence block to each variant. For variants in the intergenic region, "--nearest" option was used to identify the nearest gene with a protein-coding transcription start site (TSS) for variants.

**Replication and comparation**

As for external replication, we examined the genetic influence on TBA and ICP with two independent study cohorts (Baoan NIPT Plus cohort and BIGCS cohort). SNPs meeting the following criteria were regarded as replicated: 1) they exhibited a consistent direction of effect for lead SNPs with that of cohorts, and 2) they reached Bonferroni-corrected *P* values or passed a two-sided two-sample t-test.

The two-sided two-sample t-test was conducted to evaluate the equivalence of genetic effects on the same traits between two independent cohorts, with the following hypotheses:

$$Null\ hypothesis\ H_0: \beta_m = \beta_i$$

$$Alternative\ hypothesis\ H_1: \beta_m \neq \beta_i$$

The *T* statistic was computed as follows:

$$T = \frac{\beta_m - \beta_i}{\sqrt{\frac{S_m^2}{n_m} + \frac{S_i^2}{n_i}}} = \frac{\beta_m - \beta_i}{\sqrt{SE_m^2 + SE_i^2}} \sim t(v') \quad (1)$$

The degrees of freedom $v'$ was determined by the formula:

$$v' = \frac{(SE_m^2 + SE_i^2)^2}{\frac{SE_m^4}{n_m - 1} + \frac{SE_i^4}{n_i - 1}} \quad (2)$$

Herein, $\beta_m$ and $\beta_i$ represent the genetic effects associated with the same TBA and ICP traits for two cohorts, respectively. $S_m^2$ and $S_i^2$ denote sample variance, whereas $SE_m$ and $SE_i$ stand for estimated standard errors. It is established that the *T* statistic in equation (1) follows a *t*-distribution with a degree of freedom $v'$. To address potential inequality between $S_m^2$ and $S_i^2$ , the adjusted $v'$ was employed, computed according to formula (2).

If the lead SNP did not exist external GWAS summary datasets, the proxy SNP of the lead SNP with LD $R^2$ greater than 0.8 and existed in both data was chosen as a substitute. Proxy SNPs were queried using the LD proxy Tool (https://ldlink.nih.gov/?tab=ldproxy) through LDlink[11] (version 5.5.1) based on GRCh38 1000 Genomes Project (1000GP) genome build in East Asian (EAS) populations.

**Identification of novel locus and signals**

We identified SNPs as novel locus if no SNPs in GWAS Catalog within 1Mb block of the SNP were reported to be associated with our results. If there were SNPs has been reported before associated with the SNP in 1Mb region, LDpair (https://ldlink.nih.gov/?tab=ldpair) was used to calculate the linkage disequilibrium $R^2$ with lead SNP. If lead SNPs identified in our study were novel SNP with $R^2 < 0.2$ seem as a novel signal in a known locus.

**Colocalization analysis**

Colocalization evaluates the posterior probabilities of five mutually exclusive hypotheses: 1) $H_0$: no association of any variant in the region with either trait; 2) $H_1$:

association with first trait but not the second; 3) $H_2$: association with second trait but not the first; 4) $H_3$: associated with both traits but have two independent causal variants and 5) $H_4$: associated with both traits and shared one causal variant[12]. Colocalization analysis has been originally designed for testing two sets of associations measured on different individuals. While, previous study has confirmed by simulation that the results running it on the same individuals appear robust to correlated errors[13].

Here, we utilized PP4 (posterior probability that there exists a single causal variant common to both traits) $\geq 0.75$ and PP4/PP3 $\geq 3$ to identify colocalization between the GWAS and GWAS signals[12].

**Modern DNA-based selection test**

The high-coverage 1000 Genomes Project phased whole-genome sequencing (WGS) panel (http://ftp.1000genomes.ebi.ac.uk/vol1/ftp/data_collections/1000G_2504_high_coverage/working/20201028_3202_phased/), comprising 3,202 individuals from 26 worldwide populations, was employed for evolutionary analysis. Ancestral allele information was obtained from 1000 Genomes Project (http://ftp.1000genomes.ebi.ac.uk/vol1/ftp/release/20130502/). Python package "CrossMap.py" (https://crossmap.readthedocs.io/en/latest/)[14] was utilized to change chromosome positions from hg37 to hg38. The sample HG01783 was excluded as it belongs to both European and African ancestry. SNPs with minor allele frequencies (MAF) < 0.01 in all five super-populations and genetic variants without ancestral information were excluded. In total, 18,221,282 SNPs were used for the natural analysis.

For SFS-based tests, genetic diversity (π), Tajima's D[15], and Fay and Wu's H[16] were calculated using Perl scripts from a previously published paper[17]. These three statistics were computed with a sliding-window approach (window size = 5 kb and moving step = 1 kb). Statistical significance for these three statistics were evaluated using the genome-wide empirical distribution. Based on allele frequency differentiation and extended haplotype homozygosity, we calculated the locus-specific branch length (LSBL)[18] for the CHS population with the European population (i.e., CEU) and the African population (i.e., YRI) as reference populations. The formula used to calculate LSBL was: LSBL = $(F_{ST\ CHS\_CEU} + F_{ST\ CHS\_TRI} - F_{ST\ CEU\_YRI})/2$[19]. The 1% threshold for the whole genome was set at 0.40, and the 0.1% threshold was set at 0.58. SNPs within the top 1% were considered highly differentiated, and those within the top 0.1%were considered extremely highly differentiated.

We further integrated Haplotype Score (iHS) and the Cross Population Extended Haplotype Homozygosity (XP-EHH) test using R package rehh 2.0[20]. For iHS, we calculated the iHS value for each locus in the CHS population, considering SNPs with |iHS| > 2 as exhibiting a signal of positive selection[21]. As for XP-EHH, the score for each locus in the CHS population was calculated with CEU as the reference population, and SNPs with XP-EHH > 2 were considered to exhibit a signal of positive selection[22].

### Supplementary Figures

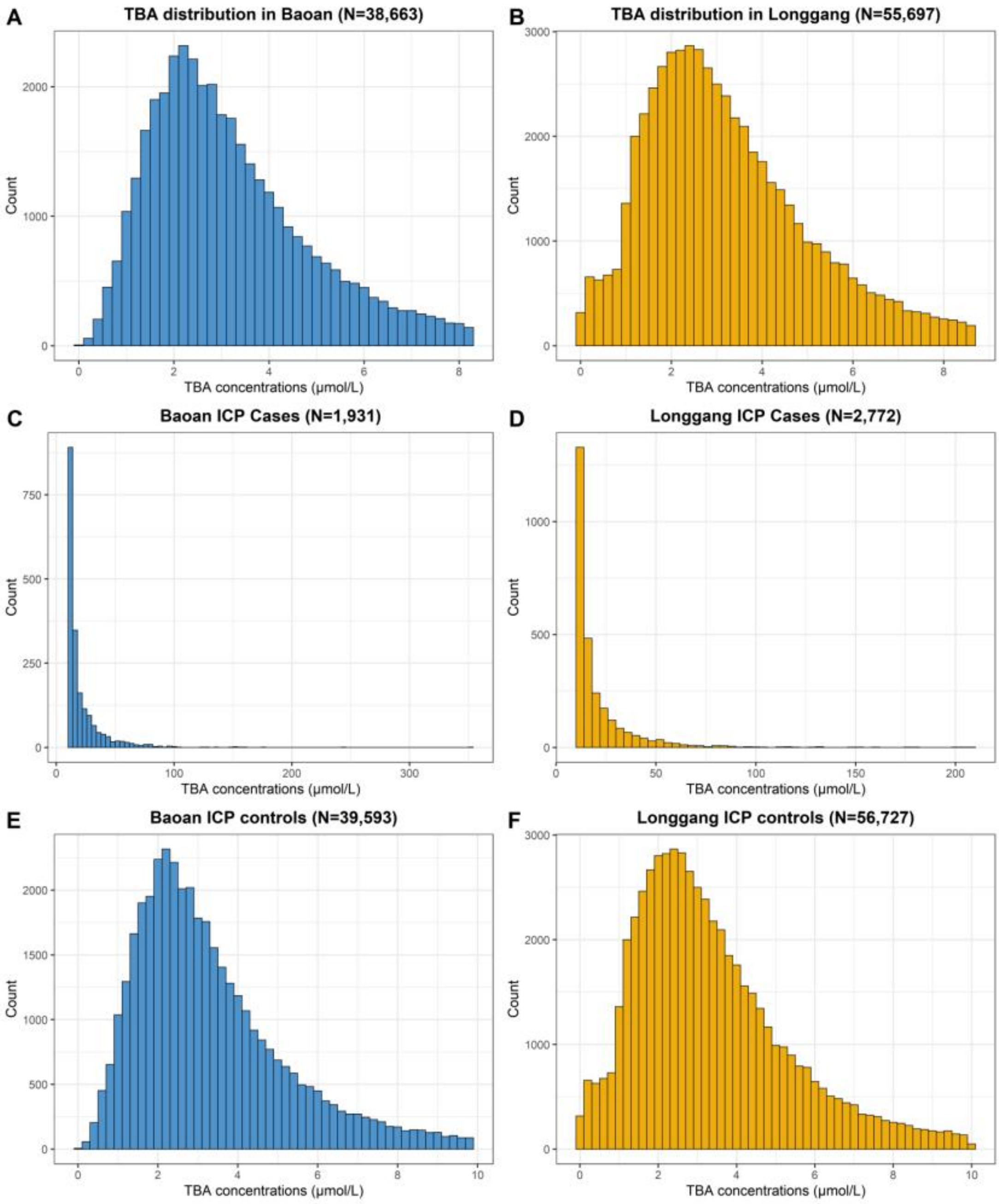

**Supplementary Fig. 1. Distribution of total bile acid concentrations in the Baoan and Longgang cohorts.**

Panel (A) and (B) depict TAB levels among normal pregnancies (TBA concentration < 10μmol/L) for each of the two hospitals. Panel (C) and (D) depict TAB levels among cases for each of the two hospitals. Panel (E) to (F) illustrate the TBA concentrations of cases and controls within the two hospitals.

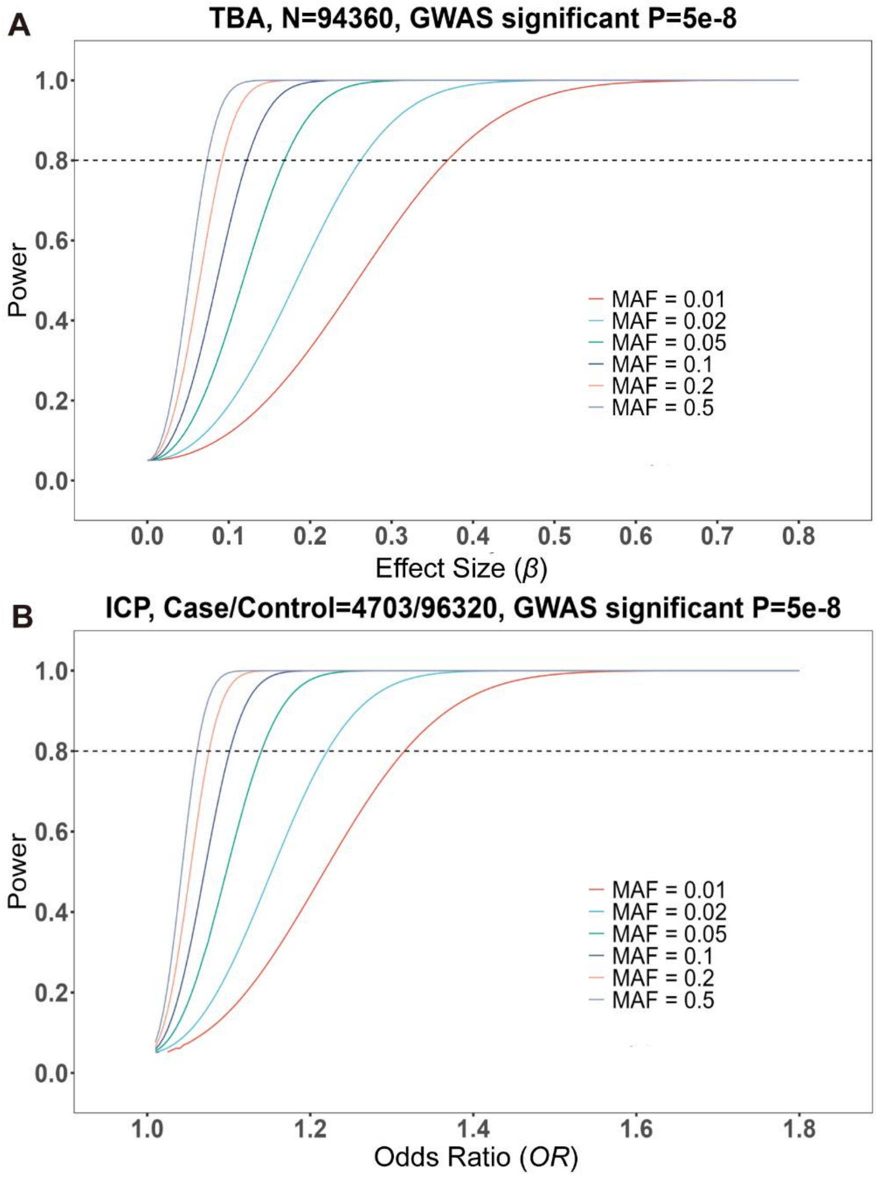

**Supplementary Fig. 2. Power calculations based on the size of the cohort for the combined meta-analysis.**

(A) Power analysis for the meta-GWAS of TBA with a sample size of 94,360. (B) Power analysis for the meta-GWAS of ICP with 4,703 cases versus 96,320 controls. The graph illustrates the ability to detect genome-wide associations at a significance threshold of *P* value < $5\times10^{-8}$ for varying odds ratio (x-axis) and minor allele frequencies (MAF). Power calculations were performed using a linear or logistic model under genetic addictively.

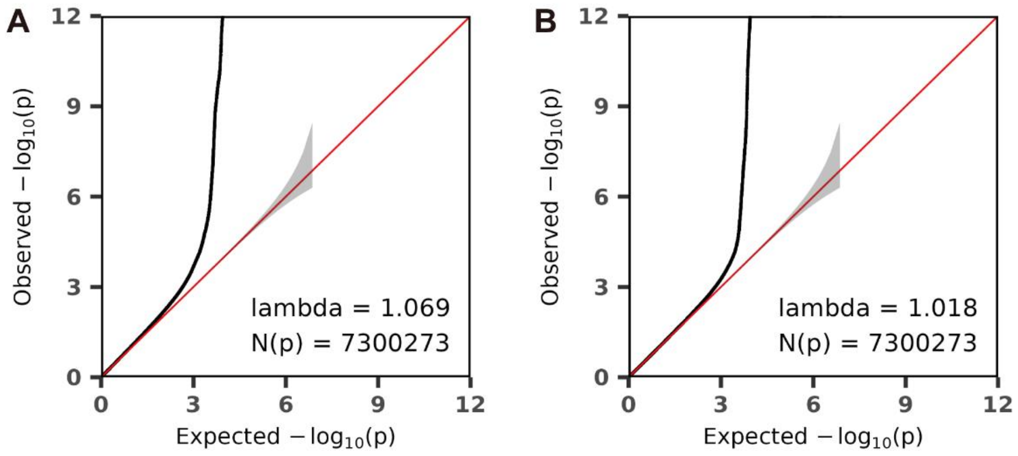

**Supplementary Fig. 3. QQ plots of TBA and ICP GWAS meta-analyses during gestational weeks 13-42.**

QQ plots for GWAS meta-analyses of (A) TBA and (B) ICP depict the relationship between observed and expected -$\log_{10}(P)$ values. The red line represents the distribution of *P* values under the null hypothesis, and the gray shaded area indicates stander errors. The genomic inflation (lambda) is shown in the QQ plots, indicating no significant population stratification.

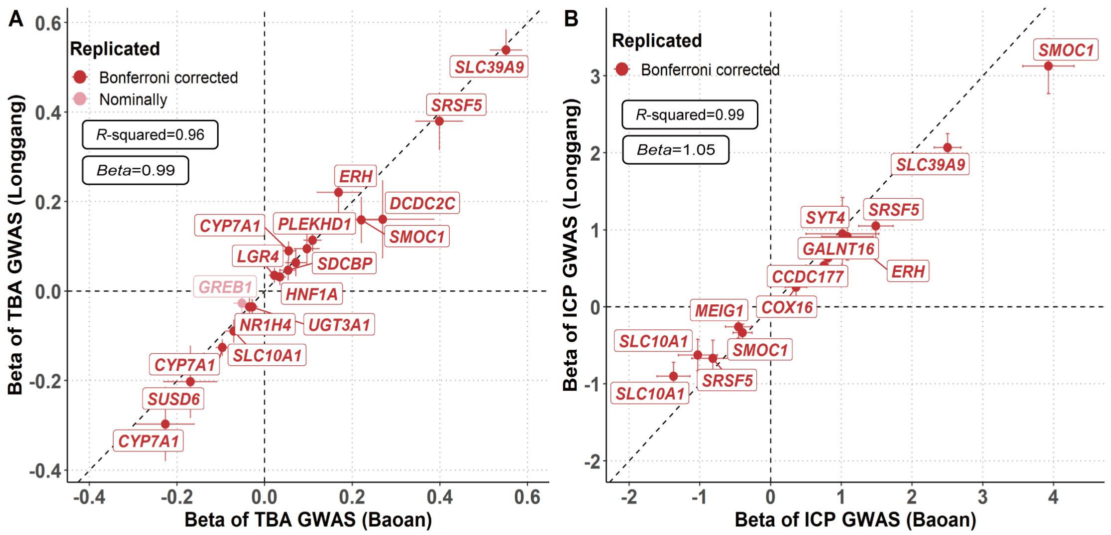

**Supplementary Fig. 4. Internal replication of TBA and ICP GWAS.**

In (A) and (B), the beta and *P* values for TBA and ICP traits were replicated in two internal cohorts (Baoan and Longgang). The x-axis shows the beta values for TBA or ICP GWAS in the Baoan cohort, while the y-axis represents the beta values for the Longgang cohort. The error bars indicate the 95% confidence interval of beta. Red points denote SNPs selected from GCTA analysis with consistent directions in beta and achieved Bonferroni corrected significant *P* values. Pink points indicate SNPs with the same beta and nominally significant *P* values. The Bonferroni significant threshold was calculated as 0.05 divided by the number of independent loci for traits. Detail data can be found in **Supplementary Table 2**.

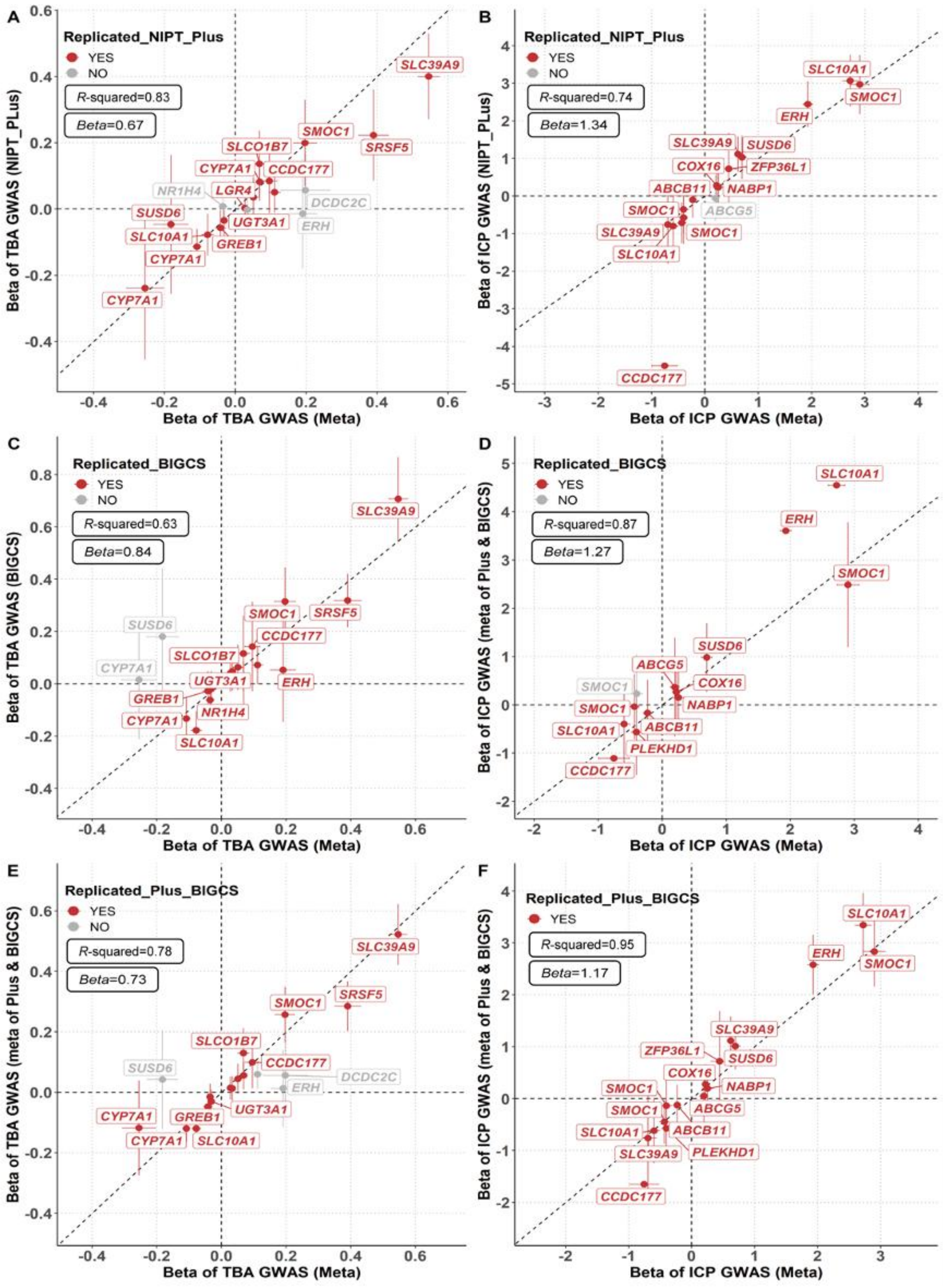

**Supplementary Fig. 5. External replication of TBA and ICP meta-GWAS using independent Chinese population cohorts.**

Panel (A) - (F) compared the meta-GWAS results of TBA and ICP against Baoan NIPT PLUS, BIGCS cohorts, and the GWAS-meta of Baoan NIPT PLUS and BIGCS cohorts. The x-axis depicts the beta values of TBA or ICP meta-GWAS, while the y-axis represents the beta values of two independent cohorts or the result of meta-analysis of these two independent cohorts. The error bars denote the 95% confidence interval of beta. Red points denote SNPs selected from GCTA that meet the following criteria: 1) exhibit consistent direction in beta, and 2) attain a significant GWAS *P* value in an external dataset after Bonferroni correction or pass the T-test with *P* value > 0.05. Grey points indicate SNPs that do not meet any of these criteria. Detailed data, excluding empty and proxy loci, are available in **Supplementary Table 2**.

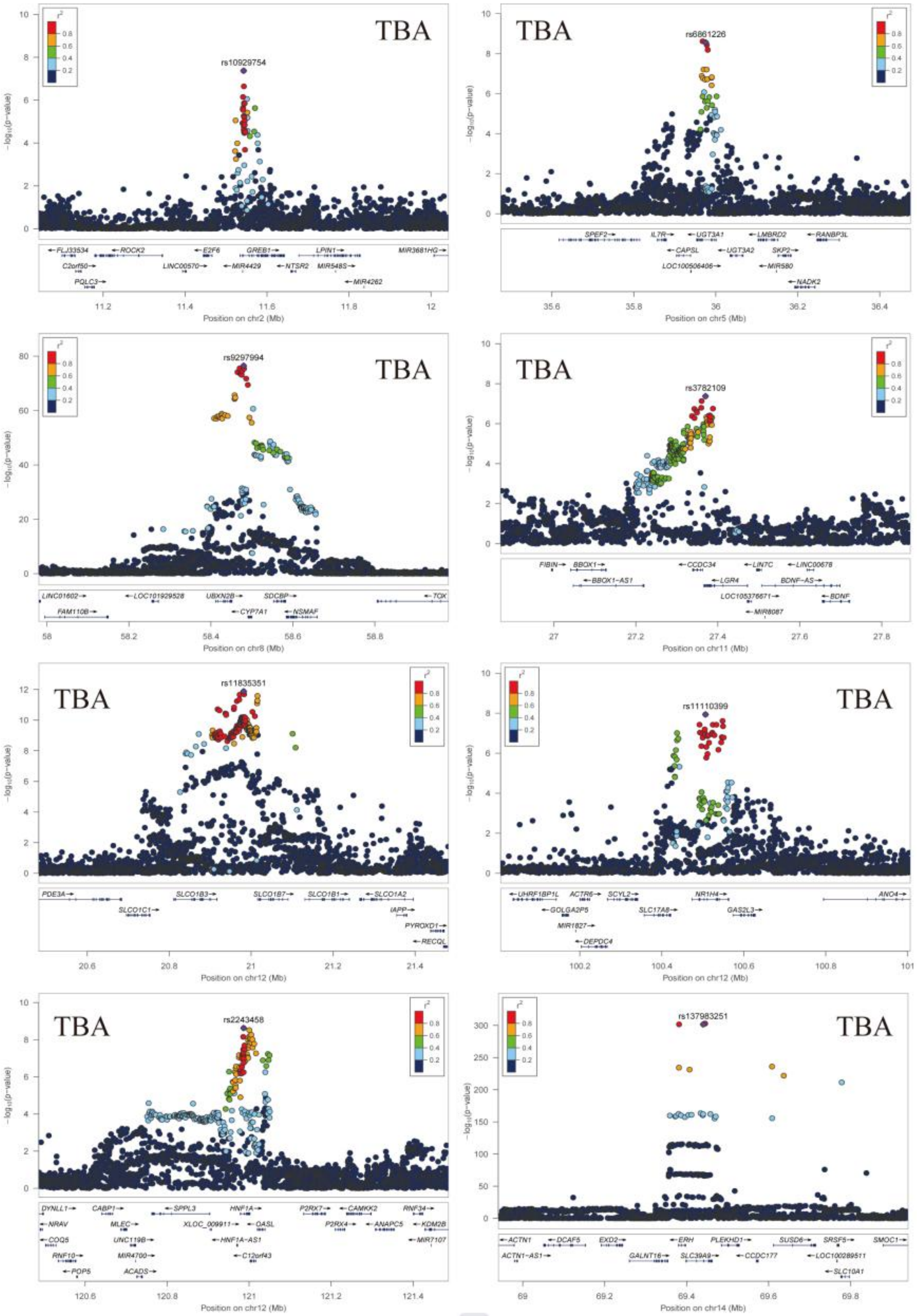

**Supplementary Fig. 6. LocusZoom plots of genome-wide significant loci associated with the TBA trait investigated in the study.**

For the eight lead SNPs associated with TBA level (excluding two SNPs, rs59789496 and rs71423384, which appear not credible), regional association and linkage disequilibrium (LD) plots were presented. These plots encompass the upstream and downstream 500kb flanking region of each lead SNP. The x-axis shows chromosome positions with respect to GRCh38, and the y-axis indicates $-\log_{10}(P)$ values for the associated tests. The purple diamond represents the lead SNP of each locus, while other SNPs are color-coded based on their LD ($r^2$) with the lead SNP. The plots were generated using LocusZoom software.

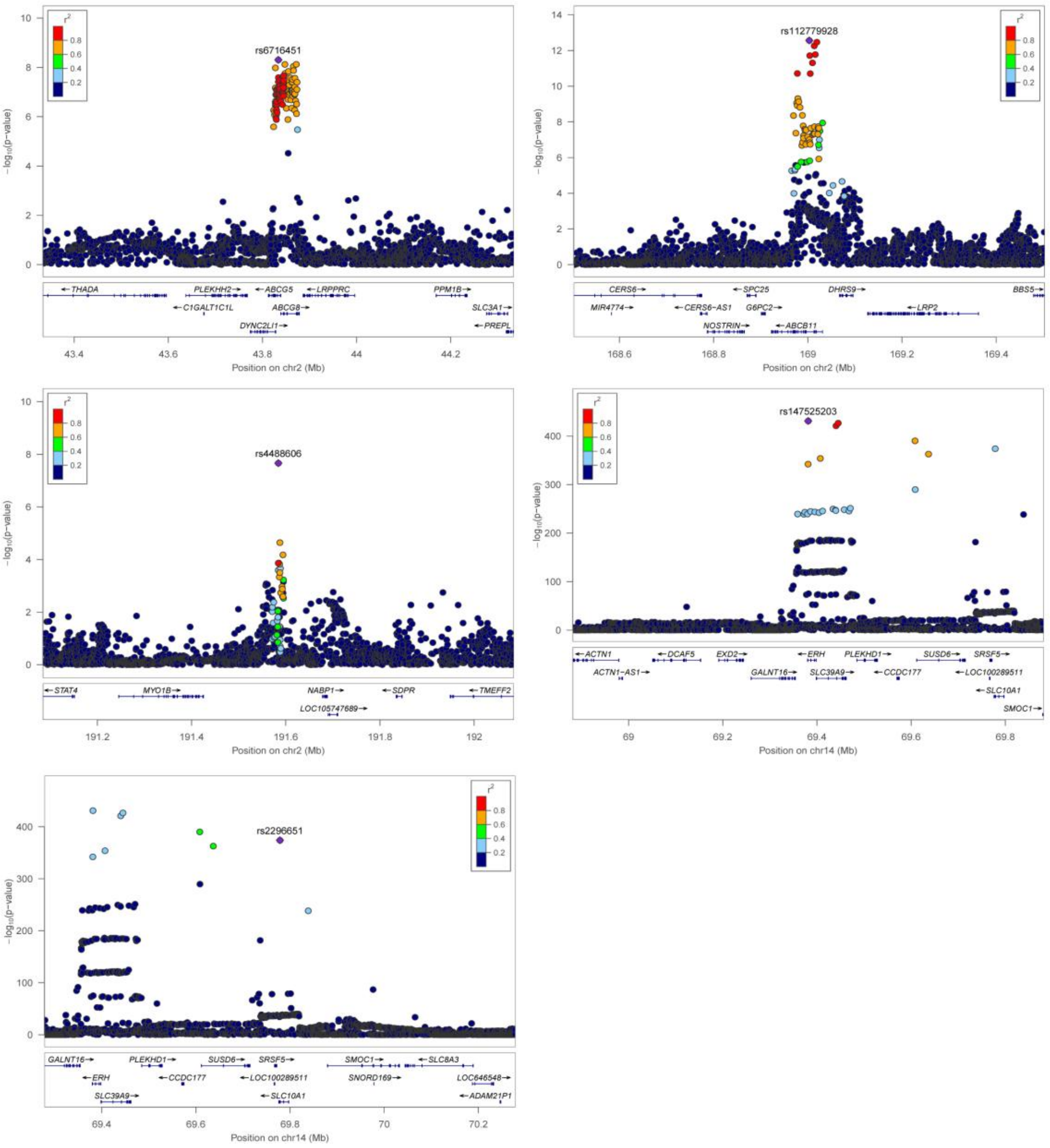

**Supplementary Fig. 7. LocusZoom plot of genome-wide significant loci associated with ICP.**

For the five lead SNPs associated with ICP (exclude two SNP, rs74573797 and rs1951510, which appear not credible), regional association and linkage disequilibrium (LD) plots were presented. These plots encompass the upstream and downstream 500kb flanking region of each lead SNP. The x-axis displays chromosome positions with respect to GRCh38, while the y-axis indicates $-\log_{10}(P)$ values for the associated tests. The purple diamond represents the lead SNP for each locus, and other SNPs are color-coded based on their LD $r^2$ with the lead SNP. Plots were generated using LocusZoom software.

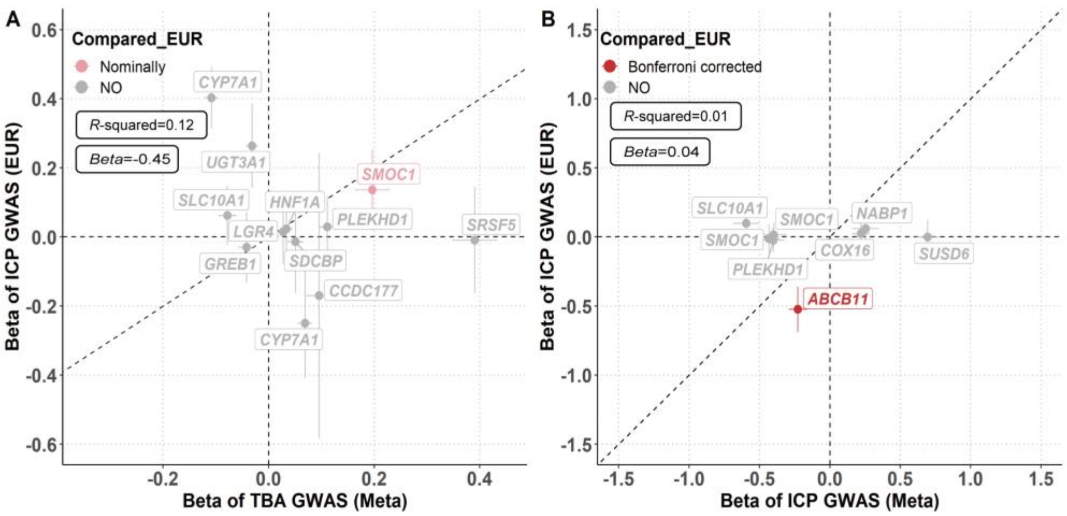

**Supplementary Fig. 8. Comparison of effect sizes between TBA and ICP GWAS with European ICP GWAS results.**

Panel (A) and (B) compared the beta and *P* values of TBA and ICP with the previously published meta-GWAS results of European population. The x-axis shows the beta values of TBA or ICP meta-GWAS, while the y-axis illustrates the beta values of the European ICP meta-GWAS result. The error bars represent the 95% confidence interval of beta. Grey points highlight instances with different directions of beta and/or *P* values > 0.05. Detail data, excluding empty and proxy loci, are available in **Supplementary Table 2**.

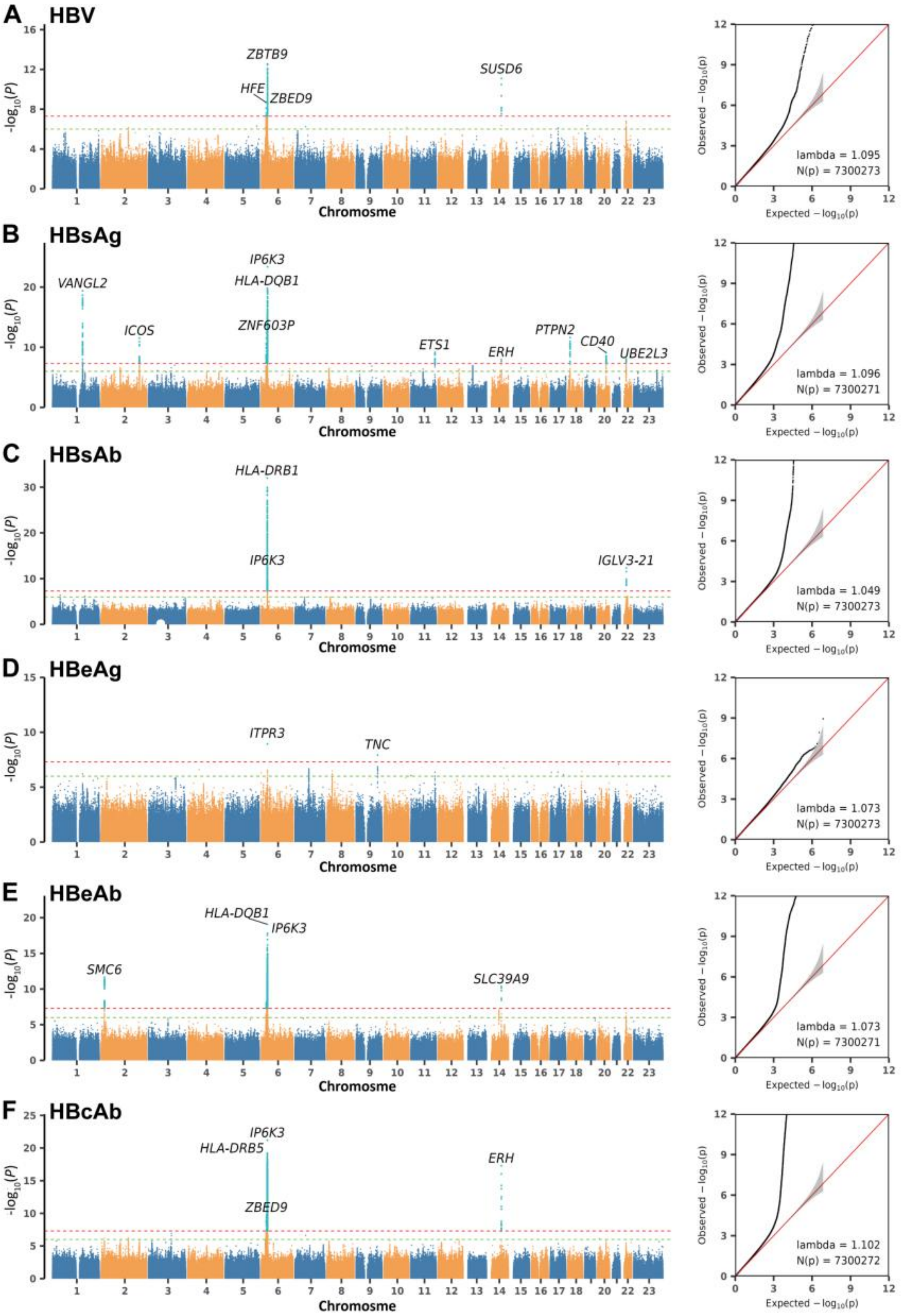

**Supplementary Fig. 9. Genome-wide association study of HBV and its antigens & antibodies during pregnancy.**

Panel (A) to (F) present the GWAS results and QQ plots of Hepatitis B virus (HBV), Hepatitis B surface antigen (HBsAg), Hepatitis B surface antibody (HBsAb),

Hepatitis B e antigen (HBeAg), Hepatitis B e antibody (HBeAb), and Hepatitis B core antibody (HBcAb) respectively. Horizontal lines delineate the genome-wide significance ($P < 5\times10^{-8}$, red line) and suggestive significance ($P < 5\times10^{-6}$, green line) thresholds. QQ plots elucidate the relationship between observed and expected $-\log_{10}(P)$ values from the GWAS meta-analysis of traits, which indicate the absence of significant population stratification. The sample size for HBV infection, as per medical records, is 44,432, while for HBsAg, HBsAb, HBeAg, HBeAb and HBcAb, it ranges from 94,441 to 94,462.

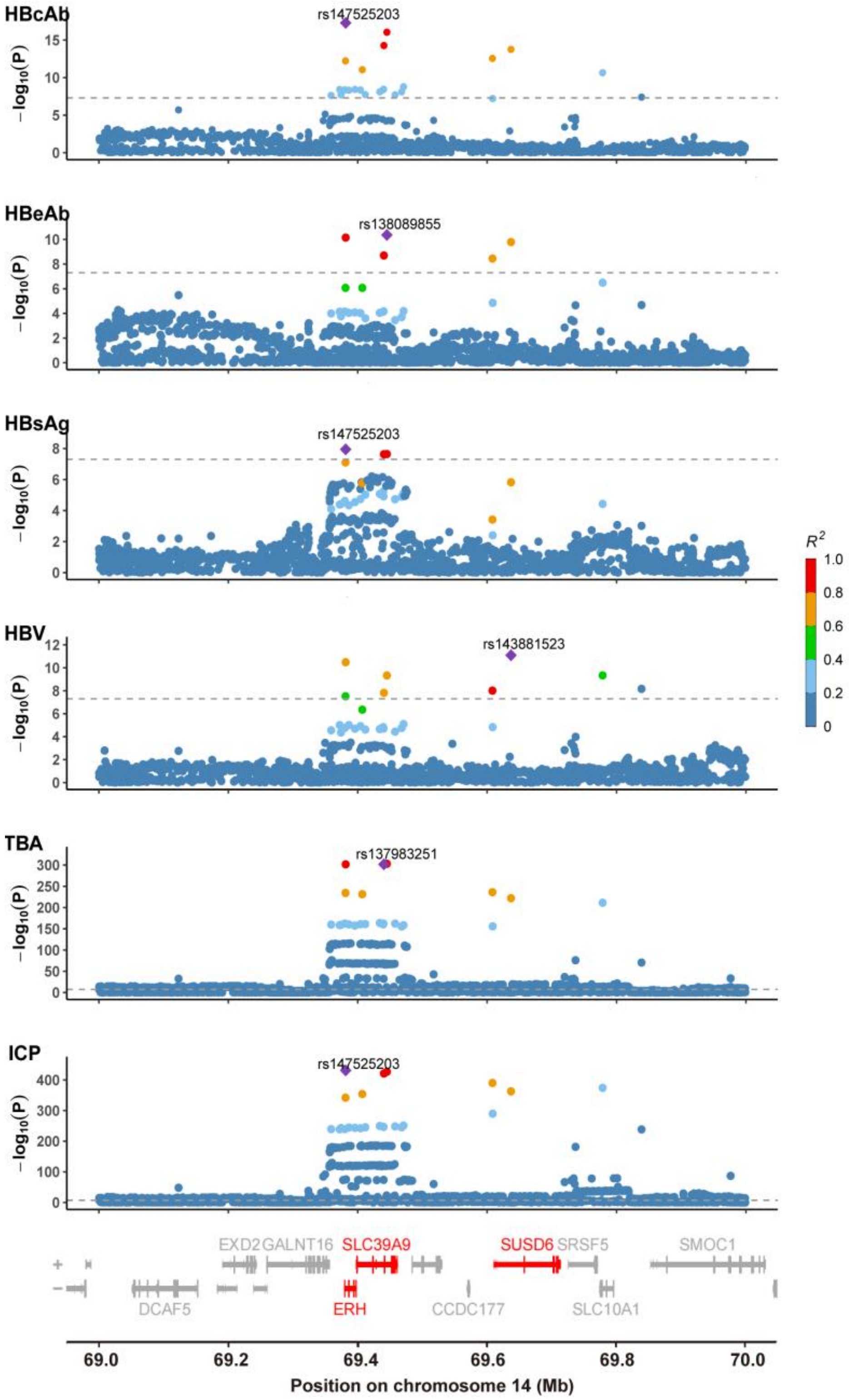

**Supplementary Fig. 10. Stacked LocusZoom plot of TBA, ICP and HBV related traits.**

Shown is a Stacked LocusZoom plot depicting TBA, ICP and HBV related phenotypes with genome-wide associations in the chromosome14 spanning 69.0-70.0

Mb region. The x-axis shows the chromosome position based on GRCh38, while the y-axis shows -$\log_{10}(P)$ values for the associated tests. Genes linked to the lead SNPs are highlighted in red, and others are marked in grey. The SNP with lowest *P* value in the locus is indicated by a purple diamond. The remaining SNPs in the region are color-coded based on their $R^2$ with the lead SNP. $R^2$ were calculated using the BIGCS reference panel. Notably, all four lead SNPs exhibit high linkage disequilibrium.

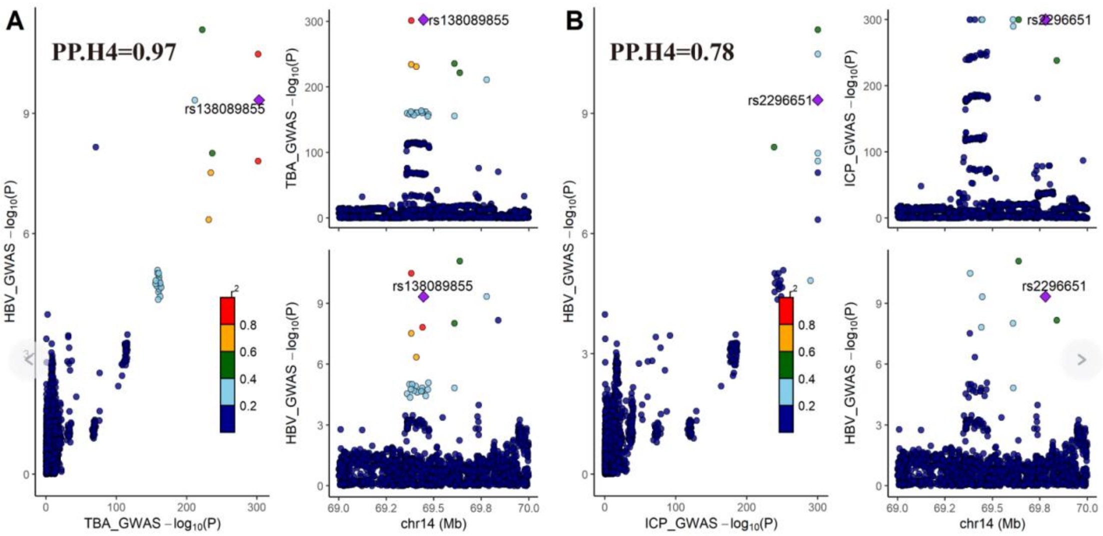

**Supplementary Fig. 11. LocusCompare plot of GWAS-GWAS colocalization between TBA & ICP and HBV.**

The x-axis shows the chromosome position based on GRCh38 and the y-axis shows $-\log_{10}(P)$ values for the associated tests. The purple diamond represents the shared SNP of (A) TBA and HBV, (B) ICP and HBV. The remaining SNPs in the region are color-coded based on $R^2$ with the lead SNP. $R^2$ were calculated with 1000GP EAS population as reference panel.

A

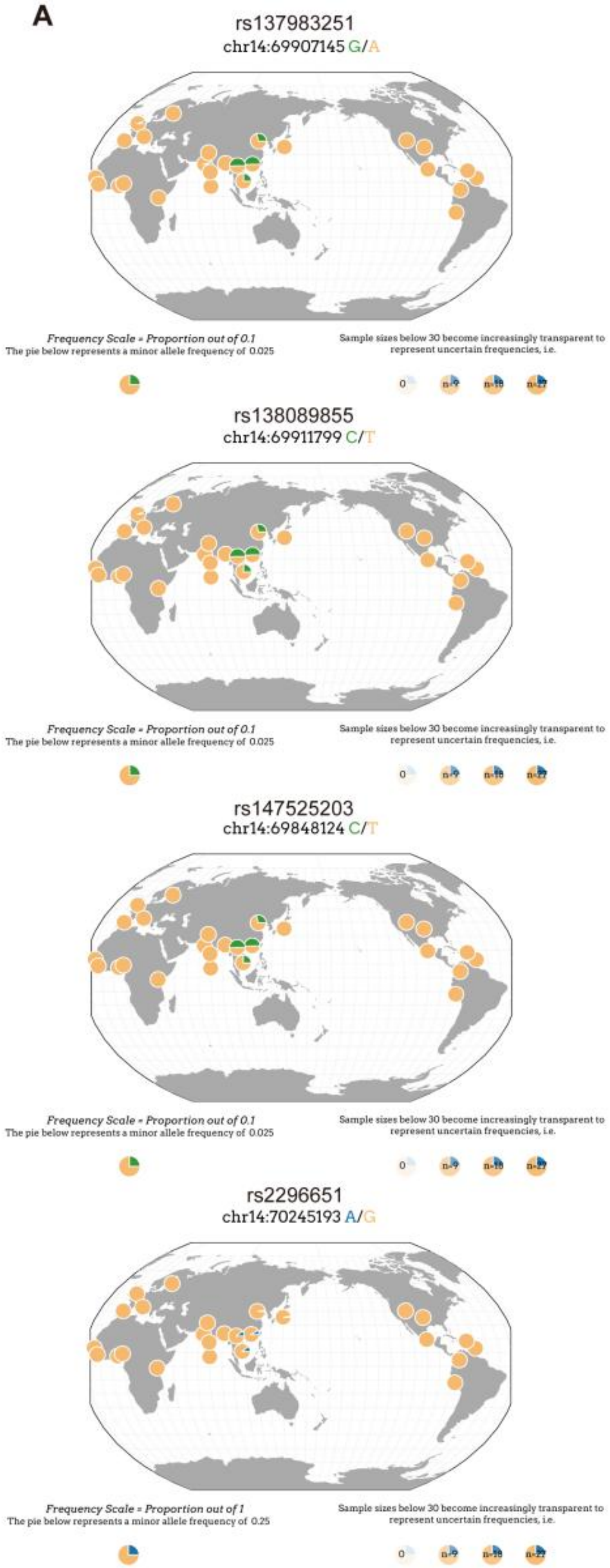

B

rs147525203 chr14:69848124

| rs137983251 chr14:69907145 | C | T | | |
|---|---|---|---|---|
| A | 0 | 979 | 979 | (0.971) |
| G | 29 | 0 | 29 | (0.029) |
| | 29 (0.029) | 979 (0.971) | 1008 | |

| Haplotypes | Statistics |
|---|---|
| A_T: 979 (0.971) | D': 1.0 |
| G_C: 29 (0.029) | $R^2$: 1.0 |
| A_C: 0 (0.0) | Chi-sq: 1008.0 |
| G_T: 0 (0.0) | p-value: <0.0001 |

rs137983251(A) allele is correlated with rs147525203(T) allele
rs137983251(G) allele is correlated with rs147525203(C) allele

rs138089855 chr14:69445082

| rs137983251 chr14:69440428 | C | T | | |
|---|---|---|---|---|
| A | 0 | 979 | 979 | (0.971) |
| G | 29 | 0 | 29 | (0.029) |
| | 29 (0.029) | 979 (0.971) | 1008 | |

| Haplotypes | Statistics |
|---|---|
| A_T: 979 (0.971) | D': 1.0 |
| G_C: 29 (0.029) | $R^2$: 1.0 |
| A_C: 0 (0.0) | Chi-sq: 1008.0 |
| G_T: 0 (0.0) | p-value: <0.0001 |

rs137983251(A) allele is correlated with rs138089855(T) allele
rs137983251(G) allele is correlated with rs138089855(C) allele

rs2296651 chr14:70245193

| rs147525203 chr14:69848124 | A | G | | |
|---|---|---|---|---|
| C | 22 | 7 | 29 | (0.029) |
| T | 50 | 929 | 979 | (0.971) |
| | 72 (0.071) | 936 (0.929) | 1008 | |

| Haplotypes | Statistics |
|---|---|
| T_G: 929 (0.922) | D': 0.7401 |
| T_A: 50 (0.05) | $R^2$: 0.2109 |
| C_A: 22 (0.022) | Chi-sq: 212.591 |
| C_G: 7 (0.007) | p-value: <0.0001 |

rs147525203(C) allele is correlated with rs2296651(A) allele
rs147525203(T) allele is correlated with rs2296651(G) allele

rs2296651 chr14:69778476

| rs138089855 chr14:69445082 | A | G | | |
|---|---|---|---|---|
| C | 22 | 7 | 29 | (0.029) |
| T | 50 | 929 | 979 | (0.971) |
| | 72 (0.071) | 936 (0.929) | 1008 | |

| Haplotypes | Statistics |
|---|---|
| T_G: 929 (0.922) | D': 0.7401 |
| T_A: 50 (0.05) | $R^2$: 0.2109 |
| C_A: 22 (0.022) | Chi-sq: 212.591 |
| C_G: 7 (0.007) | p-value: <0.0001 |

rs138089855(C) allele is correlated with rs2296651(A) allele
rs138089855(T) allele is correlated with rs2296651(G) allele

**Supplementary Fig. 12. Geographical allele frequency distribution and linkage disequilibrium (LD) of rs137983251-G, rs138089855-C, rs47525203 and rs2296651-A.**

(A) The variations in all four loci are exclusive to East Asia, displaying a distinct pattern of higher frequencies in the southern region and lower frequencies in the northern region. Geography plots were generated using the Geography of Genetic Variants Browser (https://popgen.uchicago.edu/ggv/). (B) All three loci exhibit high linkage disequilibrium (LD), with significant *P* values less than 0.0001. Plot (B) was extracted from LDpair (https://ldlink.nih.gov/?tab=ldpair).

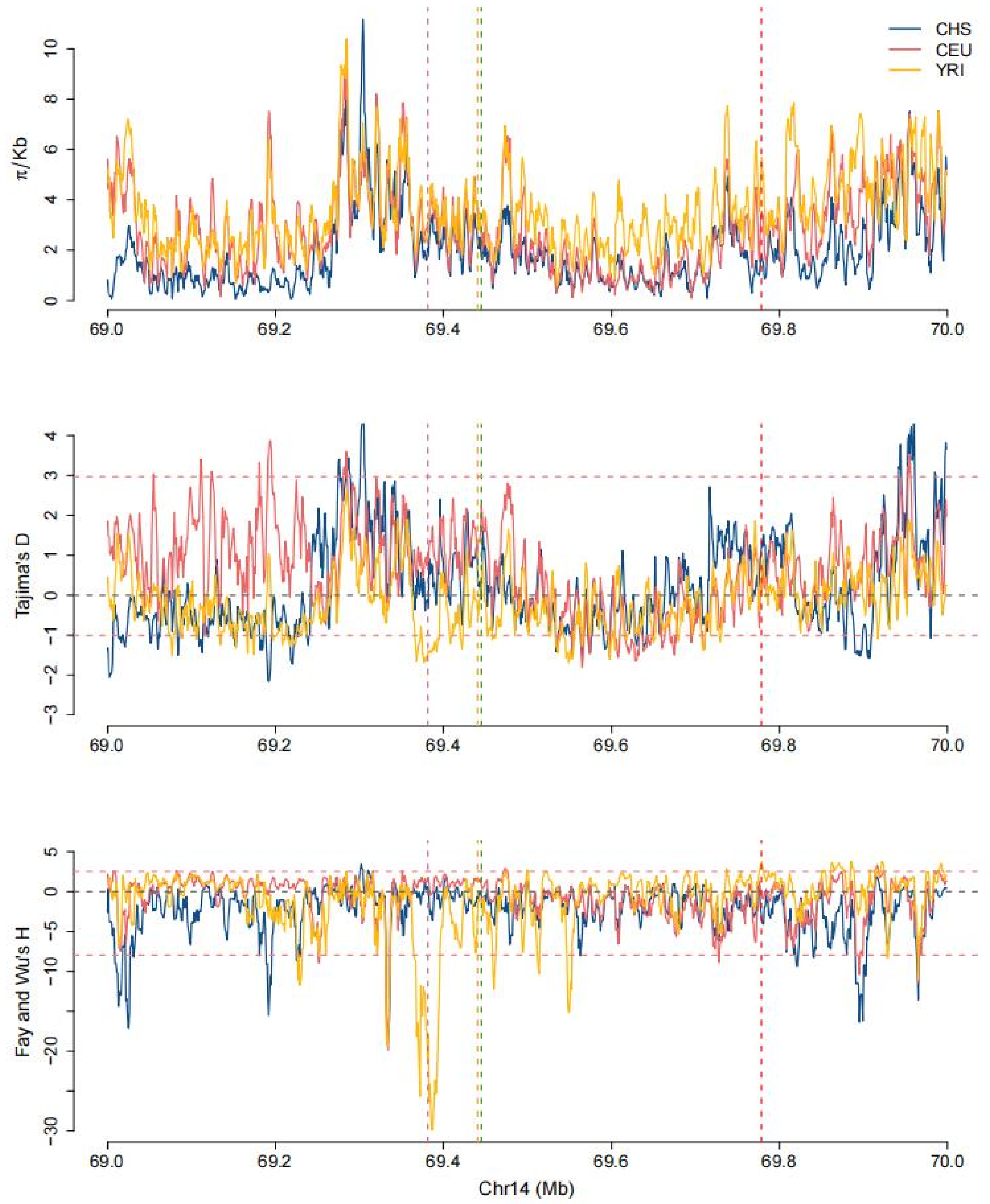

**Supplementary Fig. 13. Site Frequency Spectrum-based selection tests with modern DNA samples from 1000 Genomes Project across three populations.** Vertical dashed lines of distinct colors highlight the positions of four target Single Nucleotide Polymorphisms (SNPs): rs147525203 (palevioletred), rs137983251 (orange), rs138089855 (green) and rs2296651 (red). In the bottom two panels, a pink dashed line signifies the lowest 5% and 95% cutoffs among the three populations. Specifically, genomic regions falling below this pink dashed line achieve statistical significance across all populations. It is important to note that some genomic regions above the pink dashed line may still be significant in certain populations, as the 5% cutoffs in these populations are higher (less extreme). The populations represented are CHS (Southern Han Chinese), CEU (Utah Residents with Northern and Western European Ancestry), and YRI (Yoruba in Ibadan, Nigeria).

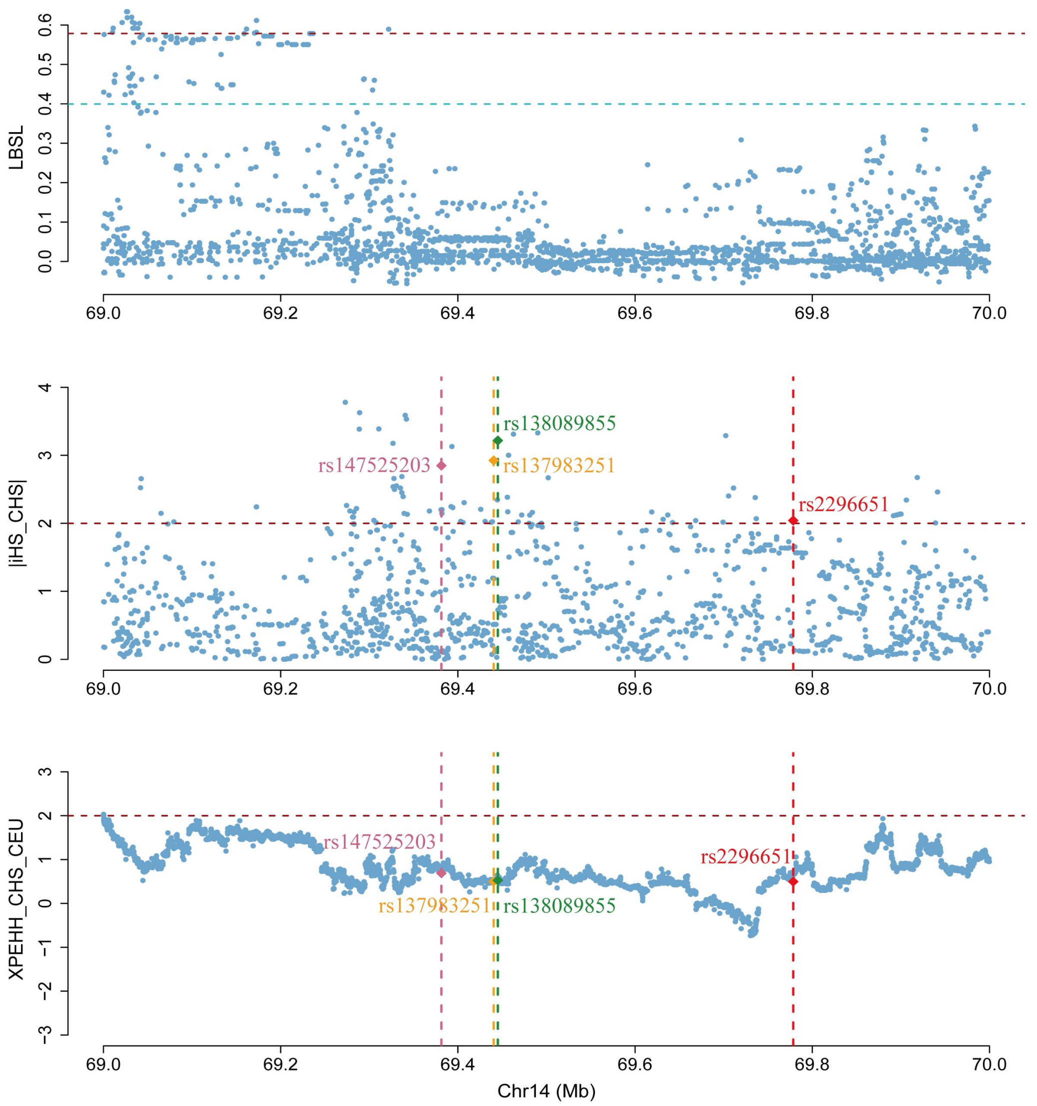

**Supplementary Fig. 14. The plot of LSBL, iHS and XP-EHH for the chromosome 14 region based on data from the 1000GP.**

The three panels represent the result of LSBL, iHS and XP-EHH, respectively. LSBL was computed for CHS with CEU using YRI as reference population. XP-EHH was calculated for CHS with CEU as the reference population. Each point on the plot represents a genetic variant. In the LSBL panel, the red and blue horizontal dashed lines denote the 0.1% and 1% threshold of the empirical distribution of the whole genome. In the iHS and XPEHH panels, the red horizontal dashed line signifies the threshold of significance. Positions for the four target SNPs are indicated with vertical dashed lines of different colors: rs147525203 (palevioletred), rs137983251 (orange), rs138089855 (green), rs2296651 (red). CHS: Southern Han Chinese; CEU: Utah

Residents (CEPH) with Northern and Western European Ancestry; YRI: Yoruba in Ibadan, Nigeria. Note that genomic portion from chr14:69.4-69.8M is missing in LSBL results due to high frequency of zero loci in the CEU population in this region.

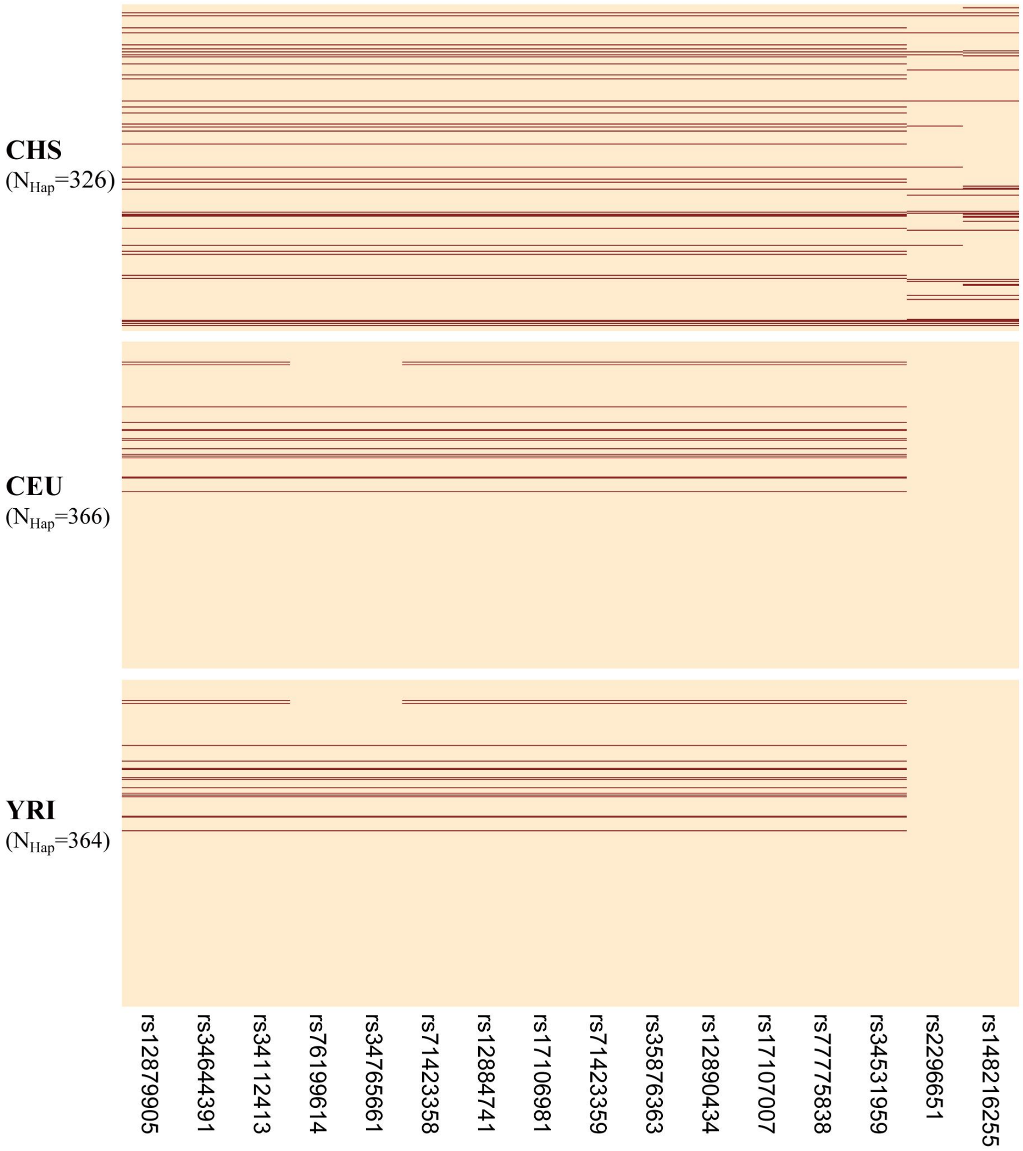

**Supplementary Fig. 15. Haplotype structure of the ICP risk 14q24.1 locus in CHS, CEU and YRI populations.**

Each horizontal line in the plot represents a distinct haplotype, and the composition of each haplotype, consisting of 16 SNPs, is presented below. The organization is organized by population (CHS, CEU, and YRI). In the plot, the blanchedalmond color is used to signify the allele in its ancestral state, while the brown color indicates a derived state.

### Supplementary Table Legends

**Supplementary Table 1**. Baseline characteristics and TBA concentrations of participants.

**Supplementary Table 2.** Genome-wide significant signals for TBA & ICP, statistics from internal and external replication and comparison with the European population

**Supplementary Table 3.** Genome-wide significant loci of TBA & ICP and internal replication.

**Supplementary Table 4.** Genome-wide significant loci of TBA & ICP and external replication in two independent cohorts in China.

**Supplementary Table 5.** Pathway and gene ontology enrichment analyses for the TBA associated gene loci (A) and the ICP associated gene loci (B)

**Supplementary Table 6.** Meta-GWAS discoveries compared with European effect estimates

**Supplementary Table 7.** GWAS-GWAS colocalization of TBA & ICP with HBV.

**Supplementary Table 8.** Frequencies of rs2296651 in the Holocene (>10,000BP) age, visualized in Fig. 3A.

**Supplementary Table 9.** Frequencies of rs2296651 in the Neolithic (10,000~3,000BP) age, visualized in Fig. 3B.

**Supplementary Table 10.** Frequencies of rs2296651 in the Historic (<3,000) age, visualized in Fig. 3C.

**Supplementary Table 11.** Frequencies of rs2296651 in the Present-day populations, visualized in Fig. 3D.

**Supplementary Table 12.** Haplotype of the lead SNPs with high Linkage disequilibrium (LD) on chromosome 14 in the 1000GP.
